# GWAS of Chronic Dizziness in the Elderly Identifies Novel Loci Implicating *MLLT10, BPTF, LINC01225*, and *ROS1*

**DOI:** 10.1101/2022.12.14.22283471

**Authors:** Royce Clifford, Daniel Munro, Daniel Dochtermann, Poornima Devineni, Saiju Pyarajan, Million Veteran Program, Francesca Talese, Abraham A. Palmer, Pejman Mohammadi, Rick Friedman

**Affiliations:** Department of Otolaryngology-Head and Neck Surgery. University of California San Diego, La Jolla, CA, USA; Research Department, Veteran Administration Hospitals, San Diego, USA; Department of Psychiatry, University of California San Diego, La Jolla, USA; Dept. of Integrative Structural and Computational Biology, Scripps Research, La Jolla, USA; Million Veteran Program, Veterans Administrations Hospitals, Boston, USA; Institute for Genomic Medicine, University of California San Diego, La Jolla, USA

**Keywords:** Dizziness, vertigo, genome-wide association study, geriatrics

## Abstract

**Background:** Chronic age-related dizziness can arise from dysfunction of the vestibulocochlear system, an elegant neuroanatomical group of pathways that mediates human perception of linear acceleration, gravity, and angular head motion. Studies indicates that 27-46% of chronic imbalance is genetically inherited, nevertheless, underlying genes leading to chronic imbalance remain unknown.

**Methods:** The Million Veteran Program comprises over 900,000 diverse-ancestry participants. Cases required two diagnoses of dizziness at least six months apart, excluding acute vertiginous syndromes, ataxias, syncope, and traumatic brain injury. Genome-wide association studies (GWAS’) were performed as separate logistic regressions on Europeans, African Americans, and those of Hispanic ancestry, followed by trans-ancestry meta-analysis. Downstream analysis included case-case-GWAS, fine-mapping, probabilistic colocalization of significant variants and genes with eQTLs, and functional analysis of significant hits.

**Results:** The final cohort consisted of 50,339 cases and 366,900 controls. Two significant loci were identified in Europeans, another in the Hispanic population, and two additional loci in trans-ancestry meta-analysis. Fine mapping revealed credible sets of intronic single nucleotide polymorphisms in genes including *MLLT10* - a histone methyl transferase cofactor, *BPTF* - a subunit of a nucleosome remodeling complex implicated in neurodevelopment, *LINC01224* - affecting transcription of *ZNF91*, a repressor of retrotransposons, and *ROS1* – a proto-oncogene receptor tyrosine kinase.

**Conclusion:** Balance dysfunction can lead to catastrophic outcomes, including falls, injury, and death in the elderly. By removing acute vertiginous syndromes and non-cochlear disorders to focus on vestibulocochlear age-related dizziness, findings suggest genomic candidates for further study and ultimate treatment of this common neurologic disease.

**KEY MESSAGES:** *What is already known on this topic:* The vestibule of the cochlea is a neuroanatomic structure mediating balance, and chronic imbalance in the elderly is a large predictor of falls and their associated morbidities and mortalities. Chronic vestibular balance is 27% - 46% heritable, however, underlying genes are unknown.

*What this study adds:* In a genome-wide association study, we identified novel single nucleotide polymorphisms and genes associated with chronic dizziness in the elderly.

*How this study might affect research, practice or policy:* Ascertaining the physiologic/genetic architecture in the cochleovestibular system will aid in future treatment where drug development can target specifically genes related to imbalance. Individuals at higher genetic risk for imbalance can be provided more focused preventive vestibular therapy.

## INTRODUCTION

Dizziness arises from dysfunction of the balance system, requiring an integration of sensory input in large part from the inner ear vestibule. This complex vestibular organ is an elegant and highly conserved neuroanatomical group of pathways that mediates our ability to perceive linear acceleration, gravity, and angular head motion. Amalgamation of vestibular, visual, and proprioception input occurs in the cerebellum, with further processing in the cerebral cortex. Animal studies document significant age-related degeneration in nearly all types of vestibular tissues, including sensory end organ hair cells, nerve fibers, and Scarpa ganglion cells [1–4].

Disorders of the vestibular system are a major source of falls [5], a leading cause of death in the elderly. Epidemiologic evidence suggests that over 6 million individuals in the US experience chronic disequilibrium that can lead to falls [6]. The incidence of dizziness increases from 22% between 65 and 69 years of age to over 40% after the age of 80 [7, 8] and there is an abundance of literature concerning associated comorbidities, including increased falls, depression, social isolation, fear, and overall functional decline [7, 9]. Thus, while falls can have multiple causes, dizziness is a strong predictor of morbidity and mortality.

Younger patients present acutely with vertigo or a whirling sensation emanating from stimuli within the three semicircular canals, utricle, and saccule in the inner ear. In contrast, as the vestibular system undergoes age-related degeneration [10] complaints are less precise, often described as an instability, dizziness, swaying, or an insecure gait, making diagnosis, treatment, and phenotyping more difficult [11, 12].

Although dizziness in the elderly encompasses multiple systems and connections, here our aim is to focus on the inner ear’s genomic role in chronic imbalance by selectively defining inclusion and exclusion criteria in the electronic health record (EHR). Evidence of a genetic component to age-related balance dysfunction includes a large Swedish population-based twin study, where self-estimated impaired balance was assessed as a risk for osteoporotic fractures and demonstrated an age- and sex-adjusted heritability of 27% [13]. An Australian twin study using the Lord’s Balance and Step tests found heritability of 46% for sensory balance modules and 30% for static and dynamic perturbations [14].

In this study, we report genome-wide association studies (GWAS) of Europeans (EU), Americans of African descent (AA) and Hispanic ancestries (LA) from the Million Veteran Program (MVP), then combined ancestries in meta-analysis. Downstream analysis seeks to define genes and single nucleotide polymorphisms (SNPs) in regard to colocalization with expression quantitative trait loci (eQTLs) in specific tissues, case-case GWAS to compare allele frequencies in a comparison cohort, and fine-mapping suggesting plausible causal variants.

## METHODS

### Participants and Phenotyping

Of the 819,397 subjects in MVP v_20_1 release, 648,224 had been assigned ancestry according to a harmonized ancestry and race/ethnicity genomic algorithm [15]. Chronic dizziness was assessed as a binary variable with a case defined as two diagnoses of dizziness or vertigo at least six months apart in the EHR. Excluded cases were those not genotyped, with no indication of sex, of only one diagnosis of dizziness or vertigo or those with diagnoses less than 6 months apart, and ICD diagnoses of an acute vertiginous syndromes, or dizziness unrelated to the cochlea, i.e., syncope, ataxias and any evidence of prior traumatic brain injury (TBI) (See Supplementary Methods and Table e1 for ICD codes with numbers of participants excluded). Since sample sizes for East and South Asian ancestry participants were too small to be statistically informative (< 10,000), these were excluded as well. Participants assigned to the control group carried no diagnoses related to dizziness or vertigo.

The final sample consisted of 417,239 participants including 50,339 cases and 366,900 controls in three ancestries. Participants had a mean age of 61.9 ± 14.3. The cohort consisted of 303,743 of European ancestry (72.80%); 79,388 participants of African American descent (19.02%); and 34,108 with Hispanic ancestry (8.17%), which is broadly consistent with the US military population. Males constituted 379,968 (91.06%), and 37,271 (8.93%) were female, also consistent with the make-up of the US military. Of the total, 255,559 were 2 50 years of age (61.25% of the cohort).

Since there was no directly applicable replication cohort, we compared our phenotype to a previously published meta-analysis with any diagnosis of vertigo [16]. Although this cannot be construed as a “replication” cohort, to our knowledge there were no chronic dizziness GWAS studies available in the literature for direct comparison. Nevertheless, both studies focused on cochleovestibular sources of dizziness.

### Genetic and Statistical Analysis

Genotyping in MVP has been well-described [17]. Briefly, genotyping had been carried out via a 723,305–SNV biobank array (Axiom; Affymetrix) customized for MVP to include variants of interest in multiple diverse ancestries. Imputation had been performed with Minimac 3 and the 1000 Genome Project phase 3 reference panel, using MVP Release 4 data (GrCh37). Final genotype data consisted of 96 million genetic variants. For each ancestry group, principal components of genotype were calculated using PLINK 2.0 alpha [18] (See Supplementary Material Genotyping, Imputation, and GWAS).

GWAS was performed as a logistic regression testing imputed dosages for association with chronic balance under an additive model using REGENIE v2.2 [19], including the first 10 principal components of genotype, sex, and age as covariates. GWAS was analyzed separately on each of the three ancestries (EU, AA, and LA), with 913,319 SNPs filtered at MAF > 0.01, INFO > 0.60, and HWE > 1 × 10^−03^. Two phenotypes were considered: one defined as two diagnoses 2 90 days apart on each ancestry - EU, AA, and LA ancestries (N = 427,959, cases = 55,404, controls = 372,555) and the other 2 180 days apart (N = 417,230, cases = 50,339, controls = 366,00). Since sample sizes including participants ≥ 50 years of age were sufficient only for EU, we then performed GWAS separately on this subset.

Summary statistics were filtered to minor allele frequency of > 1% and imputation information score > 0.6. For primary analyses, genome-wide significance was defined as p < 5.0 ×10^−08^. QQ Plots for each GWAS are in Figure e1. The proportion of inflation of test statistics due to the polygenic signal (rather than population stratification) was calculated with linkage disequilibrium score regression (LDSC), specifically, as 1 − (LDSC intercept −1)/(mean observed χ^2^− 1) [20]. Regional association plots were generated using LocusZoom with 400-kilobase windows around the index variant and linkage disequilibrium patterns calculated based on 1000 Genomes Project ancestries appropriate for each lead SNP (Supplementary Figures e2-e9) [21].

Summary statistics were combined in meta-analysis using METAL [22]. Results of each study were weighted proportional to the square-root of each study’s sample size. 17,735,101 variants remained after filtering with INFO > 0.6, MAF 2 0.01, and inconsistent allele labels and strands adjusted or removed. Cochran’s Q-test was implemented for each SNP.

In the Iceland/Finland/UKB/US comparison cohort (n = 48,072 and 894,541 controls) replication was considered significant at p-value < 0.00625, a Bonferroni adjustment for the eight significant lead SNPs from our GWAS.

The proportion of heritability explained by our GWAS was performed with linkage disequilibrium scores (LDSC) [23]. Pairwise genetic correlations (rg) between results on chronic imbalance, hearing [24] and tinnitus [25] were calculated using bivariate LDSC regression, with publicly available GWAS summary statistics. LDSC regressions were computed with the Ice/Fin/UKB/US replication cohort for comparison.

### Post-GWAS Analysis

Case-Case GWAS in R identified differences and directions of effect in allele frequencies between MVP and Ice/Fin/UK/US cohort (see Supplementary Methods) [26]. For significant SNPs (p-value = 5.0 × 10^−08^), output included statistically significant effect sizes reflecting direction and magnitude of effect and measured genetic distance between the two cohorts (See Supplementary Methods)[26].

Next, probabilistic colocalization between the GWAS associations (EU ancestry only) and expression quantitative trait loci analysis (eQTLs) was performed using fastENLOC to identify genes whose expression may mediate the GWAS association [27, 28]. We used the pre-computed eQTL annotations for GTEx v8 data from the fastENLOC repository, with SNPs labeled as rsIDs [29]. Approximately independent LD blocks for European ancestry from LDetect were also included as input [30]. fastENLOC was run separately on the eQTL data from each of the 49 tissues, and regional colocalization probability (RCP) per eQTL per tissue were examined for evidence of colocalization, using an RCP > 0.1 cutoff for putative causal gene contributors.

Fine mapping of European GWAS SNPs was computed with a “Sum of Single Effects” for summary statistics model using default settings in R, with posterior inclusion probabilities (PIP) set at 0.95 (See Supplemental Methods) [31]. Functional mapping and annotation of genetic associations (FUMA), version 1.4.2, was based on human genome assembly GRCh37 (hg19), with default settings unless stated otherwise [32]. Gene-based analysis was performed with the FUMA implementation of MAGMA version 1.08 [33]. Significance of genes was determined with a Bonferroni-corrected threshold of p-value = 0.05/18,914 protein coding genes = 2.644 × 10^−06^. eQTL gene mapping was included using GTEx v8 Brain with 13 tissues. 3D chromatin interaction mapping was performed to capture chromatin conformation with dorsolateral prefrontal cortex and hippocampus tissues (HiC GSE87112) [34].

PheWAS was performed using SNPs with the smallest p-value from fine-mapping at the Atlas of GWAS Summary, with significance based on Bonferroni correction for the number of phenotypes engaged by each SNP [35].

The STREGA reporting guidelines were used in this report [36].

## RESULTS

### Demographics

Chronic dizziness was associated with increased age, p-value < 0.001 (Table 1). Overall case prevalence was 12.06%, with case mean age 67.21 ± 11.67 versus control mean age 61.15 ± 14.45, range (19 to 108), p-value < 2.0 × 10^−16^. Women had a dizziness prevalence of 12.48% versus 13.67% for men (p-value < 2.0 × 10^−16^). Those of African ancestry were less likely to have a EHR diagnosis of chronic dizziness than those of European or Hispanic ancestry (OR 0.728, 95% CI 0.756, 0.700, and OR 0.875, 95% CI 0.841, 0.909, respectively), p-value < 2.0 × 10^−16^ and 1.97 × 10^−10^.

**Table 1.** Demographics.

|  |  | Cases | (% of category) | Controls | (% of category) | Total | (% of total) | *p-value* |
| --- | --- | --- | --- | --- | --- | --- | --- | --- |
| TOTAL |  | 50,339 | (12.06) | 366,900 | (87.94) | 417,239 | (100.00) |  |
| Age(SD) | | 67 | (11.73) | 61 | (14.62) | 62 | (14.28) | $< 0.001$ |
| AFR | male | 8,801 | (12.93) | 59,280 | (87.07) | 68,081 | (85.75) | $< 0.001$ |
| | female | 1,334 | (11.80) | 9,973 | (88.20) | 11,307 | (14.24) | $< 0.001$ |
| EUR | male | 33,605 | (11.95) | 247,696 | (88.05) | 281,301 | (92.92) | $< 0.001$ |
| | female | 2,968 | (13.23) | 19,474 | (86.77) | 22,442 | (7.08) | $< 0.001$ |
| EUR $\geq 50$ | | 31,675 | (12.39) | 223,884 | (87.60) | 255,559 | (100.00) | $< 0.001$ |
| HIS | male | 3,282 | (10.73) | 27,304 | (89.27) | 30,586 | (89.67) | $< 0.001$ |
| | female | 349 | (9.91) | 3,173 | (90.09) | 3,522 | (10.33) | $< 0.001$ |

The Ice/Fin/UKB/US cohort consisted of 48,072 cases and 894,541 controls aggregated from four European cohorts.[16] Case rate was reported as 5.10%. Although there have been studies on motion sickness, this was the largest study identified that was available on dizziness/vertigo for comparison [16, 36].

### Heritability and Genetic Correlation of Dizziness with Other Traits

SNP heritability for those of European descent was h2^SNP^ of 0.059 ± 0.0083 on the liability scale, assuming sample prevalence of 12% and population prevalence of 30% [7]. QQ plots identify a GCλ = 1.22127, indicative of inflation of the test statistics (Fig. e1). LDSC intercept = 1.027, thus polygenic effects account for 94.87% of observed inflation.

LDSC genetic correlation compared genomic dizziness results to other cochlear disorders in EU ancestries (Table e2). Our dizziness phenotype was moderately correlated with hearing difficulties, rg = 0.314 ±0.048 (p-value 5.77 × 10^−11^), and tinnitus, rg = 0.20 ± 0.8 (p-value = 1.51 × 10^−02^). The Ice/Fin/UKB/US cohort showed similar correlations for hearing, rg = 0.19 ± 0.049 (p-value 2.0 × 10^−4^) and tinnitus, *r*_*g*_ = 0.19 ± 0.049 (p-value 8.7 × 10^−05^).

### Genome-Wide Association Study and Replication

Results were not substantially different between diagnoses 90 days apart and 180 days apart (*r*_*g*_ = 1.00, p-value 0.0) (Supplementary Table e2). Thus, we report the results from the 2 180 days diagnosis only. GWAS of chronic dizziness on those of EU ancestry indicated two significant loci, lead hits rs10828248 on chromosome 10 (p-value 3.17 × 10^−08^) and rs7221651 on chromosome 17 (p-value 3.71 × 10^−10^) (Figure 1 and Table 2). GWAS of the Hispanic ancestry indicated a novel locus on chromosome 6, rs71717606 with p-value 4.84 × 10-10, unobserved in any of the other GWAS’. GWAS on the AA ancestry cohort revealed no significant findings. Regional association plots are seen in Figures e2-e9.

**Fig1.**
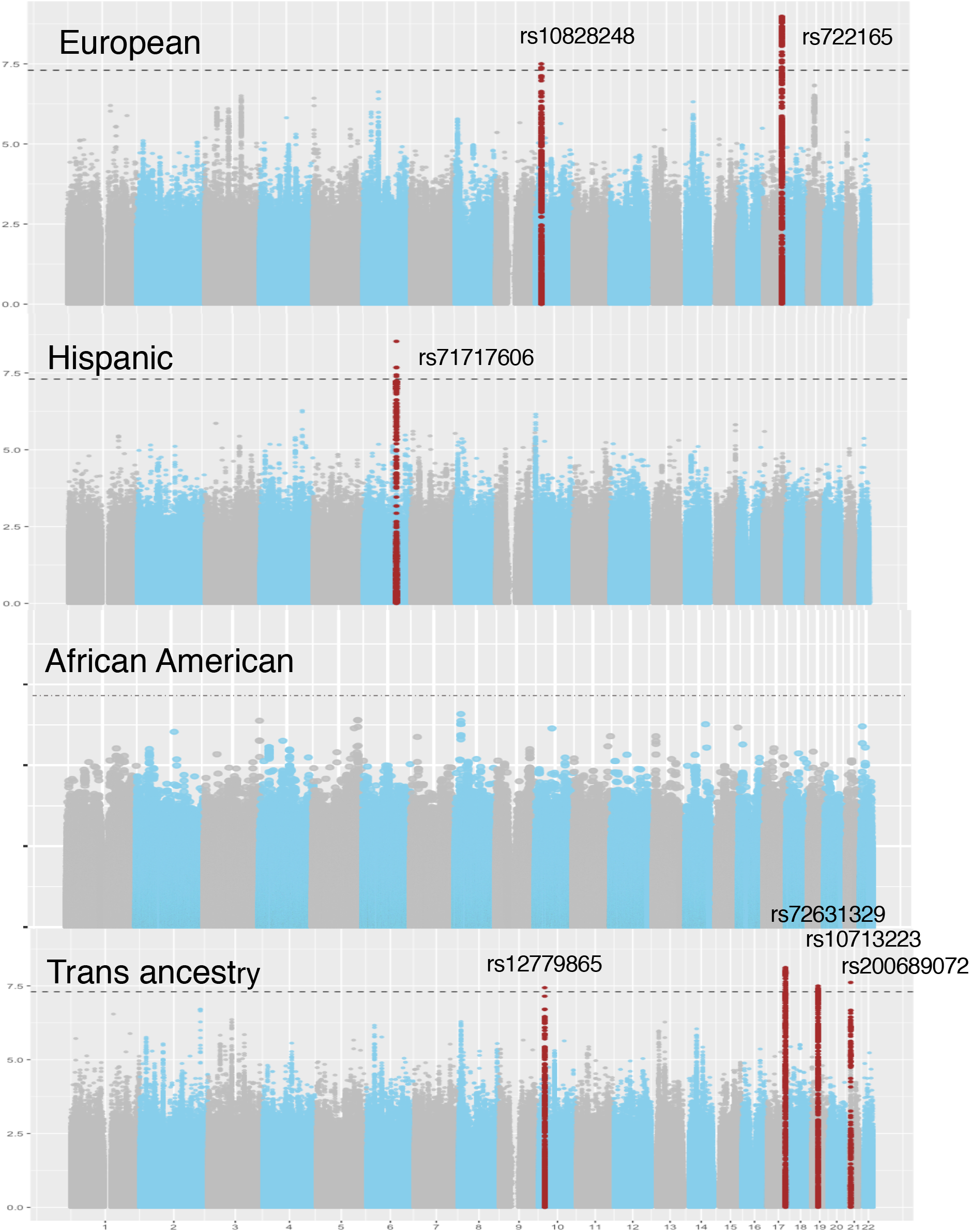
Manhattan Plot of GWAS for each ancestry, with lead SNPs indicated in each locus for which p-value < 5.0 × 10^−08^. A. GWAS of European cohort (EU). B. GWAS of Hispanic ancestry (LA). C. GWAS of African American ancestry (AA). D. Meta-analysis of European, African American, and Hispanic populations. The horizontal line represents the Bonferroni-adjusted significance threshold p-value 5 × 10^−08^. Red dots indicate single-nucleotide variations within the significant regional loci (See Supplemental Figures e2-e9 for regional plots).

**Fig2.**
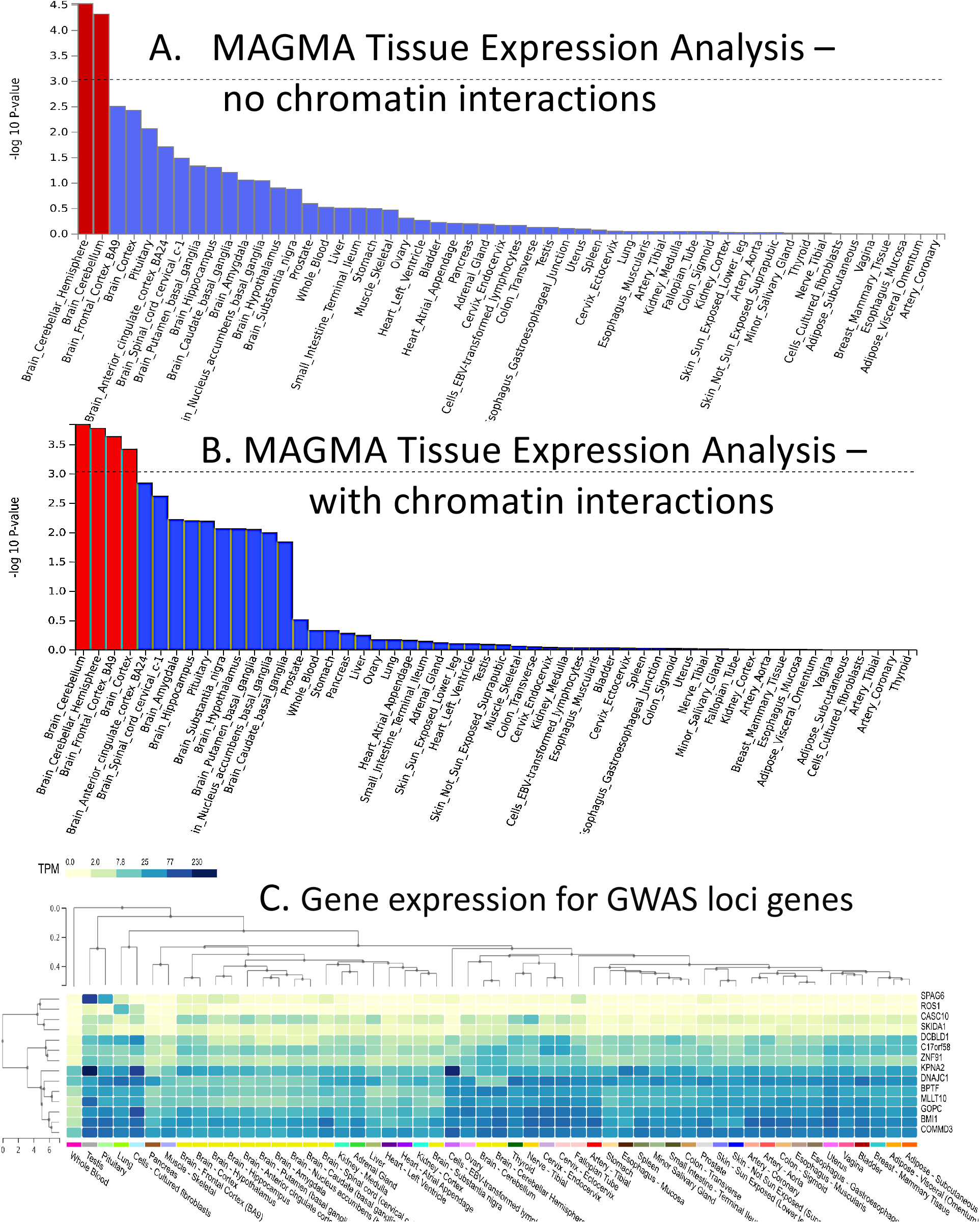
Gene enrichment comparing body tissues. A. With no chromatin enhancement, using the full distribution of SNP p-values, only cerebellum shows significant expression. Dotted line denotes significant p-value corrected for 54 tissues. B. With 3-D chromatin analysis including Hi-C and Chia_PET interchromatin analysis, other aspects of the brain are enhanced, specifically frontal cortex. C. Heatmap of gene expression for GWAS loci genes in each GTEx v8 tissue. Rows and columns are arranged by hierarchical clustering of the transcripts per million (TPM) values.

**Table 2.**
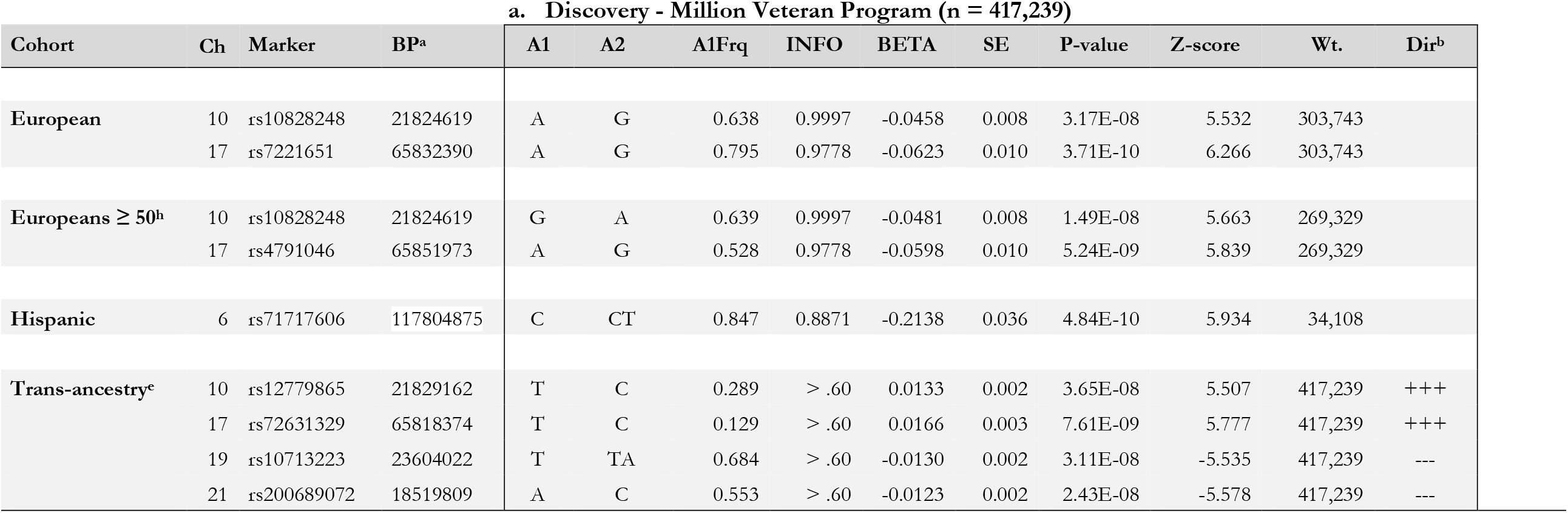
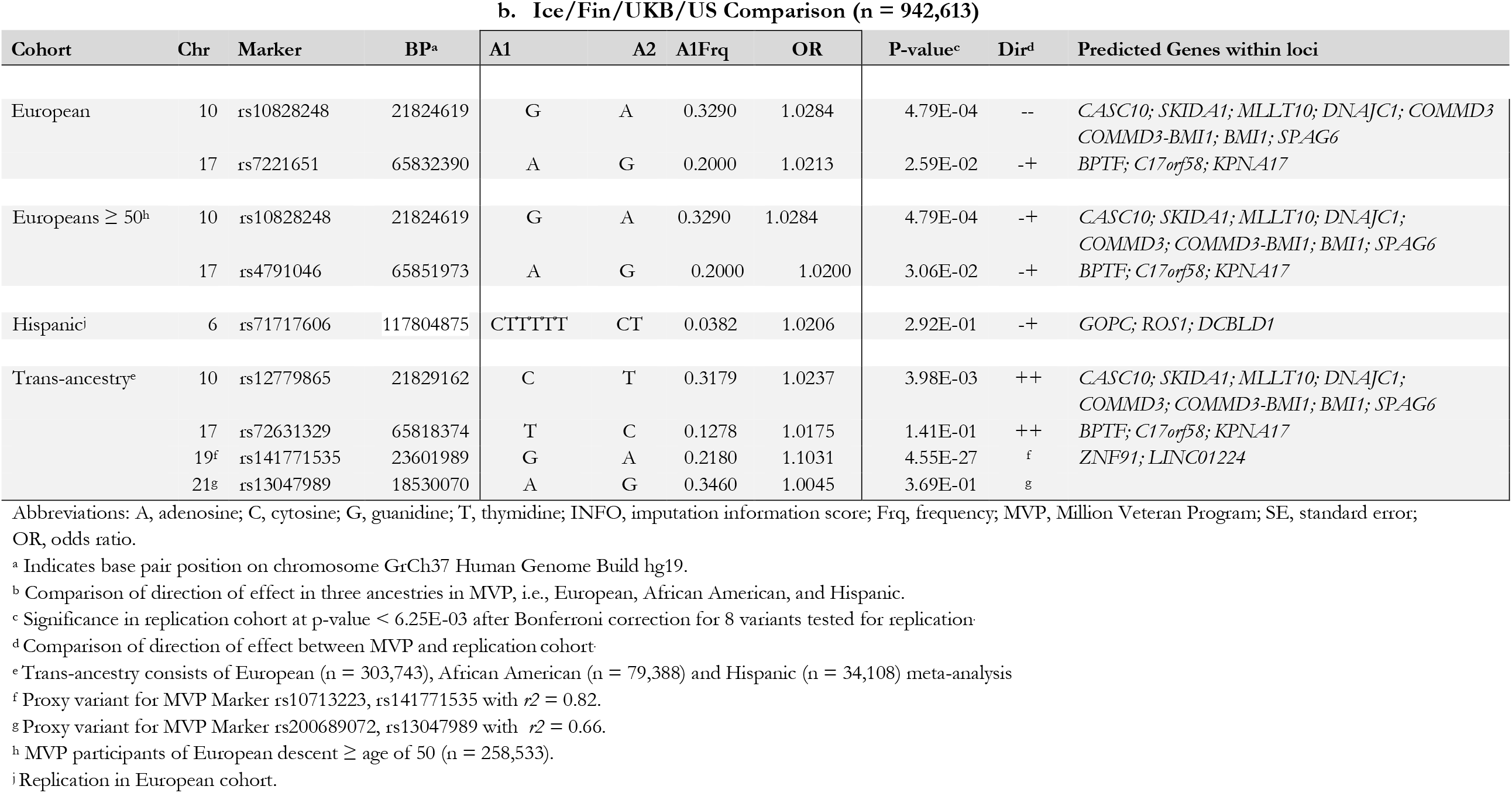
Genome-Wide Significant Loci Associated with Dizziness in the Million Veteran Program with Replication in a European Cohort.

GWAS in trans-ancestry analysis identified four significant loci at p-value < 5 × 10^−08^, including the two loci on chromosomes 10 and 17 identified in the EU GWAS. Lead SNPs in chromosomes 10 and 17 from trans-ancestry and European GWAS’ were in moderate to high linkage disequilibrium (LD). R^2^ of rs12779865/rs10828248 on chromosome 10 = 0.87 and R^2^ of rs72631329/rs7221651 on chromosome 17 = 0.54.

Comparison of GWAS on all Europeans to those 2 50 years of age yielded similar results, with no significant changes in z-scores of the identified loci on chr 10 and 17 (data not shown, Cohen’s D = 0.001 for both hits, indicative of essentially no difference in the two cohorts). Once again, lead SNPs for EU ≥ 50 were in LD with both GWAS of all EU participants and with trans-ancestry GWAS results, with an identical lead SNP on chr10. On chromosome 17, rs7221651/rs4791046 R^2^ = 0.99 for EU versus EU ≥ 50 years of age.

The MVP chronic dizziness phenotype and Ice/Fin/UKB/US vertigo phenotype demonstrated a genetic correlation of *r*_*g*_ = 0.67 ± 0.073, p-value 5.34 × 10^−20^ (Table e1). Despite differences in phenotypes, two loci were replicated in the Ice/Fin/UKB/US cohort (Table 2), lead SNPs on chromosome 10, rs12779865/rs10828248 (p-values 3.98 × 10^−03^ and 4.79 × 10^−03^, respectively) and chromosome 19 rs10713223 (p-value 1.45 × 10^−27^).

Gene set analysis identified seven mapped genes at p-value < 0.05/19,093 = 2.619 × 10^−06^ for 19,093 protein coding genes analyzed (Table 3). Two of our genes were replicated in the Ice/Fin/UKB/US cohort: *ZNF19* (p-value 6.34 × 10^−14^) and *MLLT10* (p-value 1.76 × 10^−03^) [33]. CC-GWAS [26] comparison with Ice/Fin/UKB/US indicated a sole significant difference in alleles on the chromosome 19 locus (Table e3), with lead SNP rs17473980 (p-value 1.52 × 10^−23^).

**Table 3.** Significant Genes Associated with Dizziness in Million Veteran Program.

| GWAS | GENE | Start<sup>a</sup> | Stop<sup>a</sup> | a. MVP discovery |  |  | b. Iceland/Finland/UKB/US comparison |  |  |  |
| --- | --- | --- | --- | --- | --- | --- | --- | --- | --- | --- |
|  |  |  |  | # SNPs<sup>b</sup> | # params<sup>c</sup> | P-value | #SNPs<sup>b</sup> | # param<sup>c</sup> | P-value<sup>d</sup> | Dir Dis/Rep |
| **Transancestry** | *MFHAS1* | 8640864 | 8751155 | 1016 | 101 | 1.14E-06 | 517 | 29 | 0.74379 | +- |
|  | *BPTF<sup>d</sup>* | 65821640 | 65980494 | 832 | 58 | 4.84E-08 | 280 | 13 | 0.03156 | ++ |
|  | *KPNA2<sup>d</sup>* | 66031635 | 66042958 | 78 | 20 | 1.03E-06 | 39 | 5 | 0.33328 | ++ |
|  | *ZNF91<sup>d</sup>* | 23487793 | 23578362 | 584 | 51 | 4.98E-07 | *304 | 12 | 6.14E-14 | ++ |
| **European** | *MFHAS1* | 8640864 | 8751155 | 550 | 28 | 1.02E-06 | 517 | 29 | 0.74379 | ++ |
|  | *MLLT10<sup>d</sup>* | 21823094 | 22032559 | 193 | 23 | 1.78E-06 | *208 | 32 | 0.00176 | ++ |
|  | *BPTF<sup>d</sup>* | 65821640 | 65980494 | 305 | 9 | 5.04E-10 | 280 | 13 | 0.03156 | ++ |
|  | *C17orf58<sup>d</sup>* | 65987217 | 65989765 | 3 | 1 | 2.54E-07 | 3 | 1 | 0.02915 | ++ |
|  | *KPNA2<sup>d</sup>* | 66031635 | 66042958 | 39 | 4 | 6.76E-07 | 39 | 5 | 0.33328 | ++ |
|  | *ZNF91<sup>d</sup>* | 23487793 | 23578362 | 329 | 11 | 1.58E-06 | *304 | 12 | 6.14E-14 | ++ |
| **Eur >= 50** | *SKIDA1<sup>d</sup>* | 21802407 | 21814611 | 11 | 4 | 1.92E-06 | 11 | 5 | 0.00994 | ++ |
|  | *MLLT10<sup>d</sup>* | 21823094 | 22032559 | 193 | 23 | 4.97E-07 | *208 | 32 | 0.00176 | ++ |
|  | *BPTF<sup>d</sup>* | 65821640 | 65980494 | 306 | 9 | 6.51E-09 | 280 | 13 | 0.03156 | ++ |
| **Hispanic** | *GOPC* | 117639374 | 117923691 | 1157 | 46 | 2.44E-07 | 1053 | 47 | 0.81116 | ++ |

Abbreviations: MVP, Million Veteran Program; CHR chromosome; Dir Dis/Rep, direction of effect, discovery/replication
<sup>a</sup> Start position in GrCh37 Human Genome Build 19.
<sup>b</sup> SNPs tested within gene.
<sup>c</sup> Number of variants used to compute the gene-wise statistics.
<sup>d</sup> Significance in replication cohort at $P < 6.25E-03$ for 8 genes tested.

We tested for colocalization between the European GWAS associations and eQTLs in 49 tissues assayed by the GTEx Project. Regional colocalization probabilities were below 0.1 for all eQTLs, indicating lack of evidence for gene expression in the tested tissues mediating the genetic associations with chronic dizziness. The strongest colocalizations were with *MIR1915HG* (*CASC10*) in the aorta (RCP = 0.068) and *MLLT10* in sun-exposed skin (RCP = 0.064), while all other eQTLs had RCP < 0.03.

Fine mapping was performed within significant loci using summary statistics and an LD matrix from 1000 genomes with ancestry-matched values [31]. Table 4 and Figure e10 show a list of “credible sets” with a posterior probability > 0.95 for loci identified as significant comparing *r*_*g*_ with the lead SNP in each locus. For the chromosome 6 sets, fine mapping identified SNPs within an intron of *ROS1*. The sets within chromosome 10 are within an intron of *MLLT10* and in high LD with the lead SNP. In chromosome 17, the five identified SNPs are all in equilibrium with the lead SNP, thus no conclusion as to “cause” can be made, however, they are all within introns of *BPTF*. In chromosome 19, both the lead SNP and the two identified SNPs are within a long non-coding RNA, *LINC01224*, in contrast to Ice/Fin/UKB/US GWAS, where the lead SNP was within *ZNF19*. This finding is in agreement with CC-GWAS results, which indicated a significant difference in alleles between the two studies.

**Table 4.** Fine Mapping Credible Sets.

| SNP | CHR | BP<sup>1</sup> | $r_g^2$ | ALLELE1 | ALLELE0 | A1FREQ | P | BETA | SE |
| --- | --- | --- | --- | --- | --- | --- | --- | --- | --- |
| rs72965321 | 6 | 117388218 | <sup>3</sup> | C | T | 0.944778 | 1.71E-05 | -0.22678 | 0.051611 |
| rs2883416 | 6 | 117676330 | <sup>3</sup> | C | T | 0.060377 | 4.40E-03 | 0.165584 | 0.057228 |
| rs7894565 | 10 | 21583984 | 0.7428 | T | C | 0.678874 | 2.56E-07 | -0.04545 | 0.008803 |
| rs11322024 | 10 | 21747989 | 0.7609 | TC | T | 0.379806 | 4.63E-06 | 0.045273 | 0.009866 |
| rs9899520 | 17 | 67961452 | 1.0 | A | G | 0.228443 | 2.10E-09 | 0.059577 | 0.009907 |
| rs7221651 | 17 | 67836274 | 1.0 | G | A | 0.795488 | 3.71E-10 | -0.06228 | 0.009897 |
| rs1976054 | 17 | 67837108 | 1.0 | T | G | 0.795494 | 3.73E-10 | -0.06227 | 0.009897 |
| rs7208909 | 17 | 67837430 | 1.0 | T | C | 0.795494 | 3.73E-10 | -0.06227 | 0.009897 |
| rs6504542 | 17 | 67840526 | 1.0 | T | C | 0.7955 | 3.72E-10 | -0.06228 | 0.009897 |
| rs612285 | 19 | 23421337 | 0.9075 | A | G | 0.7009 | 3.78E-08 | *-5.501 | *-5.501 |
| rs620916 | 19 | 23414408 | 0.8971 | T | C | 0.2992 | 4.21E-08 | *5.482 | *5.482 |

Abbreviations: SNP, single nucleotide polymorphism; CHR, chromosome, POS, position; $r_g^2$ , R<sup>2</sup> correlation with lead SNP in locus; ALLELE0, reference allele; ALLELE1, effect allele; SE, standard error.
<sup>1</sup> Analyzed in ancestry-specific 1000 genome sets, GrCh38.
<sup>2</sup> Linkage disequilibrium score with lead SNP in the locus. All p-values for correlations < 0.001. Data from LDLink, https://ldlink.nci.nih.gov/.
<sup>3</sup> Lead SNP not in reference sets for 1000 genomes.

### Integration with Functional Genomic Data

We examined 54 body tissues for gene enrichment using sumstats from the EU GWAS and found that although ataxias and Parkinson’s disorders were excluded from the phenotype, the cerebellum was nevertheless significantly enriched at p-value 9.4 × 10^−04^ (Figure 3 and Table e4). When reanalyzed with 3D chromatin interaction mapping based on Hi-C and Chia-PET, frontal cortex and brain cortex genes became significantly overexpressed [32, 37]. We then looked at functional analysis of lead variants for all ancestries (Table e5). All significant loci contained multiple SNPs with combined annotation dependent depletion (CADD) scores > 12.37 and RegulomeDB scores < 5, indicating that the SNP alteration is within the top 7% of deleterious substitutions and the locus contains a transcription factor binding site, a Dnase peak, and/or eQTL [38, 39].

PheWAS of > 3,300 traits and disorders for each significant SNP from fine mapping analysis showed multiple correlations (Table e6 and Figure e11 for lead SNPs). From the chr6 rs72965321 locus, the most significant hit was a correlation with skin pigmentation (p-value = 1.87 × 10-^05^) and hippocampus anisotropy in diffusion tract imaging (DTI), indicating loss of white matter tracts in the hippocampus (p-value = 8.44 × 10^−05^). The chr10 rs8904565 hit was significant for multiple areas of the brain, including anisotropy of the fornix (p-value = 3.39 × 10^−11^), posterior limb of the internal capsule (p-value 4.87 × 10^−09^), left accumbens, total brain volume, posterior corona radiata, and intra-cranial areas. Chr17 rs7221651 was significant for multiple impedance measures and other measurements of body mass (Impedance measurement of trunk fat mass, p-value 7.05 × 10^−30^. Chr19 rs612285 showed minimal significance for frequent insomnia (p-value 1.40 × 10^−04^) and menopause (p-value 2.89 × 10^−04^).

## DISCUSSION

Good balance requires integration of sensory input from cochleae, ocular system, and proprioception with output to applicable muscles of the eyes and limbs. Vestibular dysfunction increases with age and is found in 35.4% of US adults over 40 years. Imbalance carries a 12x risk of falling, leading to morbidity and mortality among older individuals [40]. Part of the reason for patients’ lack of report of vertigo is related to central compensation of vision and proprioception, and these systems degenerate with age as well [7]. Although the recently coined term “presbyvestibulopathy” has stricter diagnostic criteria, including objective clinical testing, and is uncommon in patients hospitalized with complaints of vertigo and moderate impairment [41], here we use a predominantly ambulatory cohort to identify those with the health-record diagnosis of chronic dizziness.

The differential diagnosis of dizziness includes a myriad of diagnoses, such as BPPV, unilateral vestibulopathy, orthostatic hypotension, low vision, proprioceptive impairment, and extrapyramidal disorders, among others [40]. In order to focus on sensory input in the aging cochlea, in this study we avoid “minimal phenotyping”, defined as dependence on one self-report for case definitions [42]. For chronicity, we sought participants who had at least two clinical diagnoses six months apart. Even studies that use a single clinical diagnosis rather than self-report can elicit a low sensitivity of 50% and specificity of 81%, leading to high rates of false positives in subsequent GWAS, despite examination by a clinician [42]. In order to focus further on chronic dizziness, we excluded acute and intermittent vertiginous syndromes (i.e., Meniere’s disease, BPPV, and acute vestibular neuronitis), as well as non-vestibular system gait disorders such as Parkinson’s and other ataxias. We also eliminated over 25,000 of those diagnosed with traumatic brain injury (TBI), since TBI is a strong predictor of vertigo [43].

Consistent with other studies, this GWAS reveals genetic correlation with other cochlear disorders such as hearing loss and tinnitus [24, 25]. Further, despite differences in phenotype, and despite differences in phenotype definitions, genetic correlation with Ice/Fin/UKB/US is still reasonably strong and highly significant (rg = 0.67 se 0.07, p-value 5.34 × 10^−20^), indicating that our “chronic dizziness” definition shares common genetic influences with other acute cochlear and vestibular syndromes.

In post-GWAS analysis, we elicit potential causal SNPs and genes from significant loci. All lead SNPs and significant fine-mapping SNPs identified are intronic or within long non-coding RNA. SNPs identified by fine mapping include intronic variants in the gene histone lysine methyltransferase *DOT1L* cofactor (*MLLT10*) within an enhancer region. *MLLT10* is expressed in the vestibule and cochlea [44] and is thought to prevent somatic cell reprogramming through regulation of *DOT1L*-mediated H3K79 methylation. *MLLT10* is overexpressed in dark cells of the crista ampullaris in the vestibule and mesenchymal melanocytes of the utricle that maintain the endocochlear potential important for continuous. Interestingly, *MLLT10* in pheWAS is highly correlated with skin pigmentation and the melanocytes responsible for this characteristic. Further, a SNP in high LD with the top hit rs10828248 has been associated with the genesis of meningiomas (*R*^*2*^ = 0.8403, p-value < 0.001) [45].

Fine mapping suggests intronic SNPs in the gene bromodomain PHD finger transcription factor (*BPTF*) in chromosome 17. *BPTF* is the largest subunit of the nucleosome remodeling complex NURF and has been implicated in neurodevelopmental disorders, developmental delay, and autism [46]. While pheWAS has identified predominant associations with body mass rather than neurological function, in animal studies this gene is identified in Type I and Type II hair cells of the utricle in higher abundance than in the cochlea [44].

Trans-ancestry GWAS identifies rs10713223 (p-value 3.11 × 10^−08^), an insertion/deletion variant on chromosome 19, which replicates the locus from Ice/Fin/UKB/US study. However, the comparison study notes the gene of interest to be the nearby *ZNF91* [16]. CC-GWAS for this locus is highly significant at p-value = 4.55× 10^−27^, indicating a difference in allele frequencies for the lead variants between the two studies. In contrast, our lead SNP and fine-mapping analysis identified a credible set of SNPs within *LINC01224* [47]. *LINC01224* affects the transcription of *ZNF91* by regulation of long-range interactions between the *ZNF91* enhancer and promoter [47]. *ZNF91* is a transcription factor specifically required to repress primate specific SINE-VNTR-Alu retrotransposons [48]. The interplay of regulatory elements in this locus is yet to be determined.

Another novel variant, rs71717606 (p-value 4.84 × 10^−10^) is an insertion/deletion within an intron of *Discoidin, CUB*, and *LCCL* domain-containing protein (*DCBLD1*) on chromosome 6. *DCBLD1* is a transmembrane receptor involved in development [49]. The top variant rs71717606 lies within a promoter site. In contrast, fine mapping pointed to rs72965321within an intron of an adjacent *ROS1*. eQTL analysis in FUMA reveals significant expression of ROS1 in the frontal cortex, with significant HiC loops to the hippocampus (p-value < 2.3 × 10^−11^ after Bonferroni correction) [32], indicative of enhancer-promoter and promoter-promoter interactions within *ROS1. ROS1* is expressed solely in Scarpa’s ganglion and has not been identified within the vestibular portion of the cochlea [44]. To our knowledge, this is the first variant specific to those of Hispanic ancestry.

Although our phenotype was crafted to eliminate causes of chronic dizziness outside the vestibular, nevertheless, gene enrichment identifies the cerebellum as significantly enriched. This demonstrates the intimate relationship of the vestibule with the cerebellum portion of the finely tuned balance system. 3D chromatin interaction mapping uncovers genetic connections to the frontal cortex as well.

Limitations of this study include the fact that our sample is predominantly male. The trans-ancestry meta-analysis over-samples from Europeans and does not include individuals of Asian ancestries. The Million Veteran Program cohort has unique battle-related environmental exposures that have yet-to-be defined interactions with genetics and the aging process. In addition, the use of electronic health records could lead to errors of omission for several reasons. First, some of the individuals in our study receive medical care outside the VA and second, patients with dizziness may perceive it to be normal for their age or be concerned with other health problems, consistent with a slightly lower prevalence of chronic dizziness identified in national surveys. Although our GWAS-eQTL colocalization analysis data did not identify any strong hits, those data do not contain expression information from the vestibule. Nevertheless, cochleovestibular animal studies indicate that these genes are all expressed within the vestibule or vestibular ganglion [44]. It is possible that human eQTL data from cochleovestibular tissues could provide functional insight into the mechanisms underlying the identified genomic signals.

In this study we have identified the first loci associated with chronic imbalance related to the vestibular system, including the first significant locus in a Hispanic population. Downstream analysis has further clarified correlation of eQTL through colocalization [27], CC-GWAS [26], fine-mapping [31], and associated physiologic and anatomic information. We have identified loci containing regulatory loci, i.e., *MLLT10* and *BPTF*, a gene identified in the vestibular ganglion (*ROS1*) and a non-coding RNA, *LINC01224*, that may control its downstream gene *BPTF* within the same locus. Ascertaining the physiologic/genetic architecture in the cochleovestibular system will aid in future treatment where drug development can target specifically genes related to imbalance in the elderly. Individuals at higher genetic risk for imbalance can be provided more focused vestibular therapy. Future investigations with larger and more gender-balanced cohorts will build on these results to clarify the role of the vestibular system in the genomics of chronic dizziness in the elderly.

## Data Availability

All data produced in the present study are available upon reasonable request to the Million Veteran Program, and will be available at dbGap upon approval for publication in a peer-reviewed journal.

## FUNDING, CONSENT, DATA AVAILABILITY

### Funding Support

This work was supported by grants from NIH R01DC020052 and Veterans Administration RR&D MVP010. Research is based on data from the Million Veteran Program, Office of Research and Development, Veterans Health Administration. This publication does not represent the views of the Department of Veteran Affairs or the United States Government.

### Consent

Prior written informed consent for use of health records and genomic data was obtained for all participants in this retrospective analysis within the Million Veteran Program. This study was approved by Central VA IRB reference E22-36, qualifying for exemption under category 4(ii).

### Data and Code Availability

Summary statistics of GWAS are available through the Database of Phenotypes and Genotypes (dbGaP) accession number phs001672. dbGaP requires an application requesting data per standard procedure.

Algorithms are described in the manuscript and supplemental materials. There was no custom code or mathematical algorithm formulated for this research, however, standard methods as described in other manuscripts as referenced were performed.

## References

1 Matheson AJ, Darlington CL, Smith PF. Further evidence for age-related deficits in human postural function. J Vestib Res 1999;9:261–4.

2 Rauch SD, Velazquez-Villaseñor L, Dimitri PS, Merchant SN. Decreasing hair cell counts in aging humans. Ann N Y Acad Sci 2001;942:220–7.

3 Jang YS, Hwang CH, Shin JY, Bae WY, Kim LS. Age-related changes on the morphology of the otoconia. Laryngoscope 2006;116:996–1001.

4 Walther LE, Westhofen M. Presbyvertigo-aging of otoconia and vestibular sensory cells. J Vestib Res 2007;17:89–92.

5 Ekvall Hansson E, Magnusson M. Vestibular asymmetry predicts falls among elderly patients with multi-sensory dizziness. BMC Geriatr 2013;13. doi:10.1186/1471-2318-13-77

6 Harun A, Semenov YR, Agrawal Y. Vestibular function and activities of daily living: analysis of the 1999 to 2004 National Health and Nutrition Examination Surveys. Gerontol Geriatr Med 2015;1:ePub.

7 Fernández L, Breinbauer HA, Delano PH. Vertigo and dizziness in the elderly. Front Neurol 2015;6:144.

8 Cutson TM. Falls in the elderly. Am Fam Physician 1994;49:149–56.

9 Teixeira AR, Wender MH, Gonçalves AK, Freitas CDLR, Santos AMPV dos, Soldera CLC. Dizziness, physical exercise, falls, and depression in adults and the elderly. Int Arch Otorhinolaryngol 2016;20:124–31.

10 Tanioka H, Tanioka S, Kaga K. Vestibular aging process from 3D physiological imaging of the membranous labyrinth. Published online First: 2020. doi:10.1038/s41598-020-66520-w

11 Wassermann A, Finn S, Axer H. Age-associated characteristics of patients with chronic dizziness and vertigo. J Geriatr Psychiatry Neurol 2022;35:580–5.

12 Müller KJ, Becker-Bense S, Strobl R, Grill E, Dieterich M. Chronic vestibular syndromes in the elderly: Presbyvestibulopathy—an isolated clinical entity? Eur J Neurol 2022;29:1825–35.

13 Wagner H, Melhus H, Pedersen NL, Michaëlsson K. Heritability of impaired balance: a nationwide cohort study in twins. Osteoporosis Int 2009;20:577–83.

14 el Haber N, Hill KD, Cassano A-MT, Paton LM, Macinnis RJ, Cui JS, Hopper JL, Wark JD. Genetic and environmental influences on variation in balance performance among female twin pairs aged 21-82 years. Am J Epidemiol 2006;164:246–56.

15 Fang H, Hui Q, Lynch J, Honerlaw J, Assimes TL, Huang J, Vujkovic M, Damrauer SM, Pyarajan S, Gaziano JM, DuVall SL, O’Donnell CJ, Cho K, Chang K-M, Wilson PWF, Tsao PS, Sun Y v., Tang H, Gaziano JM, Ramoni R, Breeling J, Chang K-M, Huang G, Muralidhar S, O’Donnell CJ, Tsao PS, Muralidhar S, Moser J, Whitbourne SB, Brewer J v., Concato J, Warren S, Argyres DP, Stephens B, Brophy MT, Humphries DE, Do N, Shayan S, Nguyen X-MT, Pyarajan S, Cho K, Hauser E, Sun Y, Zhao H, Wilson P, McArdle R, Dellitalia L, Harley J, Whittle J, Beckham J, Wells J, Gutierrez S, Gibson G, Kaminsky L, Villareal G, Kinlay S, Xu J, Hamner M, Haddock KS, Bhushan S, Iruvanti P, Godschalk M, Ballas Z, Buford M, Mastorides S, Klein J, Ratcliffe N, Florez H, Swann A, Murdoch M, Sriram P, Yeh SS, Washburn R, Jhala D, Aguayo S, Cohen D, Sharma S, Callaghan J, Oursler KA, Whooley M, Ahuja S, Gutierrez A, Schifman R, Greco J, Rauchman M, Servatius R, Oehlert M, Wallbom A, Fernando R, Morgan T, Stapley T, Sherman S, Anderson G, Sonel E, Boyko E, Meyer L, Gupta S, Fayad J, Hung A, Lichy J, Hurley R, Robey B, Striker R. Harmonizing genetic ancestry and self-identified race/ethnicity in genome-wide association studies. The American Journal of Human Genetics 2019;105:763–72.

16 Skuladottir ATh, Bjornsdottir G, Nawaz MS, Petersen H, Rognvaldsson S, Moore KHS, Olafsson PI, Magnusson SH, Bjornsdottir A, Sveinsson OA, Sigurdardottir GR, Saevarsdottir S, Ivarsdottir E v, Stefansdottir L, Gunnarsson B, Muhlestein JB, Knowlton KU, Jones DA, Nadauld LD, Hartmann AM, Rujescu D, Strupp M, Walters GB, Thorgeirsson TE, Jonsdottir I, Holm H, Thorleifsson G, Gudbjartsson DF, Sulem P, Stefansson H, Stefansson K. A genome-wide meta-analysis uncovers six sequence variants conferring risk of vertigo. Commun Biol 2021;4:1148.

17 Hunter-Zinck H, Shi Y, Li M, Gorman BR, Ji SG, Sun N, Webster T, Liem A, Hsieh P, Devineni P, Karnam P, Gong X, Radhakrishnan L, Schmidt J, Assimes TL, Huang J, Pan C, Humphries D, Brophy M, Moser J, Muralidhar S, Huang GD, Przygodzki R, Concato J, Gaziano JM, Gelernter J, O’Donnell CJ, Hauser ER, Zhao H, O’Leary TJ, Tsao PS, Pyarajan S. Genotyping array design and data quality control in the Million Veteran Program. Am J Hum Genet 2020;106:535–48.

18 Chang CC, Chow CC, Tellier LCAM, Vattikuti S, Purcell SM, Lee JJ. Second-generation PLINK: Rising to the challenge of larger and richer datasets. Gigascience 2015;4:1–16.

19 Mbatchou J, Barnard L, Backman J, Marcketta A, Kosmicki JA, Ziyatdinov A, Benner C, O’dushlaine C, Barber M, Boutkov B, Habegger L, Ferreira M, Baras A, Reid J, Abecasis G, Maxwell E, Marchini J. Computationally efficient whole-genome regression for quantitative and binary traits. Nat Genet 2021;53. doi:10.1038/s41588-021-00870-7

20 Bulik-Sullivan BK, Loh P-R, Finucane HK, Ripke S, Yang J, Patterson N, Daly MJ, Price AL, Neale BM. LD Score regression distinguishes confounding from polygenicity in genome-wide association studies. Nat Genet 2015;47:291–5.

21 Pruim RJ, Welch RP, Sanna S, Teslovich TM, Chines PS, Gliedt TP, Boehnke M, Abecasis GR, Willer CJ, Frishman D. LocusZoom: Regional visualization of genome-wide association scan results. In: Bioinformatics. 2011. doi:10.1093/bioinformatics/btq419

22 Willer CJ, Li Y, Abecasis GR. METAL: fast and efficient meta-analysis of genomewide association scans. Bioinformatics 2010;26:2190–1.

23 Finucane, H, Bulik-Sullivan B, Gusev A, Trynka G, Reshef Y, Neale B, Price A. Partitioning heritability by functional annotation using genome-wide association summary statistics. Nat Genet 2015;47:1228–35.

24 Wells HRR, Freidin MB, Abidin FNZ, Payton A, Dawes P, Munro KJ, Morton CC, Moore DR, Dawson SJ, Frances M. Genome-wide association study identifies 44 independent genomic loci for self-reported adult hearing difficulty in the UK Biobank cohort. Am J Hum Genet 2019;:1–15.

25 Clifford RE, Maihofer AX, Stein MB, Ryan AF, Nievergelt CM. Novel risk loci in tinnitus and causal inference with neuropsychiatric disorders among adults of european ancestry. JAMA Otolaryngol Head Neck Surg 2020;146:1015–25.

26 Peyrot WJ, Price AL. Identifying loci with different allele frequencies among cases of eight psychiatric disorders using CC-GWAS. Nat Genet 2021;53:445–54.

27 Hukku A, Pividori M, Luca F, Pique-Regi R, Im HK, Wen X. Probabilistic colocalization of genetic variants from complex and molecular traits: promise and limitations. The American Journal of Human Genetics 2021;108:25–35.

28 Pividori M, Rajagopal PS, Barbeira A, Liang Y, Melia O, Bastarache L, Park Y, Consortium Gte, Wen X, Im HK. PhenomeXcan: Mapping the genome to the phenome through the transcriptome. Sci Adv 2020;6. doi:10.1126/sciadv.aba2083

29 GTEx Consortium. The GTEx Consortium atlas of genetic regulatory effects across human tissues. Science (1979) 2020;369:1318–30.

30 Berisa T, Pickrell JK. Approximately independent linkage disequilibrium blocks in human populations. Bioinformatics 2015:btv546.

31 Zou Y, Carbonetto P, Wang G, Stephens M. Fine-mapping from summary data with the ‘Sum of Single Effects’ model. Published Online First: 2022. doi:10.1371/journal.pgen.1010299

32 Watanabe K, Taskesen E, van Bochoven A, Posthuma D. Functional mapping and annotation of genetic associations with FUMA. Nat Commun 2017;8:1826.

33 de Leeuw CA, Mooij JM, Heskes T, Posthuma D. MAGMA: generalized gene-set analysis of GWAS data. PLoS Comput Biol 2015;11:e1004219.

34 Lieberman-Aiden E, van Berkum NL, Williams L, Imakaev M, Ragoczy T, Telling A, Amit I, Lajoie BR, Sabo PJ, Dorschner MO, Sandstrom R, Bernstein B, Bender MA, Groudine M, Gnirke A, Stamatoyannopoulos J, Mirny LA, Lander ES, Dekker J. Comprehensive Mapping of Long-Range Interactions Reveals Folding Principles of the Human Genome. Science (1979) 2009;326:289–93.

35 Watanabe K, Stringer S, Frei O, Umicevic Mirkov M, de Leeuw C, Polderman TJC, van der Sluis S, Andreassen OA, Neale BM, Posthuma D. A global overview of pleiotropy and genetic architecture in complex traits. Nat Genet 2019;51:1339–48.

36 Little J, Higgins JP, Ioannidis JP, Moher D, Gagnon F, von Elm E, Khoury MJ, Cohen B, Davey-Smith G, Grimshaw J, Scheet P, Gwinn M, Williamson RE, Zou GY, Hutchings K, Johnson CY, Tait V, Wiens M, Golding J, van Duijn C, McLaughlin J, Paterson A, Wells G, Fortier I, Freedman M, Zecevic M, King R, Infante-Rivard C, Stewart A, Birkett N; STrengthening the REporting of Genetic Association Studies. STrengthening the REporting of Genetic Association Studies (STREGA): An Extension of the STROBE Statement.

37 Hromatka BS, Tung JY, Kiefer AK, Do CB, Hinds DA, Eriksson N. Genetic variants associated with motion sickness point to roles for inner ear development, neurological processes and glucose homeostasis. Hum Mol Genet 2015;24:2700–8.

38 Vietri Rudan M, Barrington C, Henderson S, Ernst C, Odom DT, Tanay A, Hadjur S. Comparative Hi-C reveals that CTCF underlies evolution of chromosomal domain architecture. Cell Rep 2015;10:1297–309.

39 Rentzsch P, Witten D, Cooper GM, Shendure J, Kircher M. CADD: predicting the deleteriousness of variants throughout the human genome. Nucleic Acids Res 2019;47. doi:10.1093/nar/gky1016

40 Agrawal Y, Carey JP, della Santina CC, Schubert MC, Minor LB. Disorders of balance and vestibular function in US adults: data from the National Health and Nutrition Examination Survey, 2001-2004. Arch Intern Med 2009;169:938–44.

41 Agrawal Y, van de Berg R, Wuyts F, Walther L, Magnusson M, Oh E, Sharpe M, Strupp M. Presbyvestibulopathy: Diagnostic criteria consensus document of the classification committee of the Bárány Society. Journal of Vestibular Research 2019;29:161–70.

42 Cai N, Revez JA, Adams MJ, Andlauer TFM, Breen G, Byrne EM, Clarke T-K, Forstner AJ, Grabe HJ, Hamilton SP, Levinson DF, Lewis CM, Lewis G, Martin NG, Milaneschi Y, Mors O, Müller-Myhsok B, Penninx BWJH, Perlis RH, Pistis G, Potash JB, Preisig M, Shi J, Smoller JW, Streit F, Tiemeier H, Uher R, van der Auwera S, Viktorin A, Weissman MM, MDD Working Group of the Psychiatric Genomics Consortium, Kendler KS, Flint J. Minimal phenotyping yields genome-wide association signals of low specificity for major depression. Nat Genet 2020;52:437–47.

43 Sessoms PH, Gottshall KR, Sturdy J, Viirre E. Head stabilization measurements as a potential evaluation tool for comparison of persons with TBI and vestibular dysfunction with healthy controls. Mil Med 2015;180:135–42.

44 Orvis J, Gottfried B, Kancherla J, Adkins RS, Song Y, Dror AA, Olley D, Rose K, Chrysostomou E, Kelly MC, Milon B, Matern MS, Azaiez H, Herb B, Colantuoni C, Carter RL, Ament SA, Kelley MW, White O, Bravo HC, Mahurkar A, Hertzano R. gEAR: Gene Expression Analysis Resource portal for community-driven, multi-omic data exploration. Nat Methods 2021;18:843–4.

45 Walsh KM, Zhang C, Calvocoressi L, Hansen HM, Berchuck A, Schildkraut JM, Bondy ML, Wrensch M, Wiemels JL, Claus EB. Pleiotropic MLLT10 variation confers risk of meningioma and estrogen-mediated cancers. Neurooncol Adv 2022;4. doi:10.1093/noajnl/vdac044

46 Stankiewicz P, Khan TN, Szafranski P, Slattery L, Streff H, Vetrini F, Bernstein JA, Brown CW, Rosenfeld JA, Rednam S, Scollon S, Bergstrom KL, Parsons DW, Plon SE, Vieira MW, Quaio CRDC, Baratela WAR, Acosta Guio JC, Armstrong R, Mehta SG, Rump P, Pfundt R, Lewandowski R, Fernandes EM, Shinde DN, Tang S, Hoyer J, Zweier C, Reis A, Bacino CA, Xiao R, Breman AM, Smith JL, Katsanis N, Bostwick B, Popp B, Davis EE, Yang Y. Haploinsufficiency of the chromatin remodeler BPTF causes syndromic developmental and speech delay, postnatal microcephaly, and dysmorphic features. The American Journal of Human Genetics 2017;101:503–15.

47 Fu W, Zhao J, Hu W, Dai L, Jiang Z, Zhong S, Deng B, Huang Y, Wu W, Yin J. LINC01224/ZNF91 promote stem cell-like properties and drive radioresistance in non-small cell lung cancer. Cancer Manag Res 2021;Volume 13:5671–81.

48 He D, Wu D, Muller S, Wang L, Saha P, Ahanger SH, Liu SJ, Cui M, Hong SJ, Jain M, Olson HE, Akeson M, Costello JF, Diaz A, Lim DA. miRNA-independent function of long noncoding pri-miRNA loci. Proceedings of the National Academy of Sciences 2021;118. doi:10.1073/pnas.2017562118

49 Schmoker AM, Weinert JL, Markwood JM, Albretsen KS, Lunde ML, Weir ME, Ebert AM, Hinkle KL, Ballif BA. FYN and ABL regulate the interaction networks of the DCBLD receptor family. Molecular & Cellular Proteomics 2020;19:1586–601.

50 Hunt SE, McLaren W, Gil L, Thormann A, Schuilenburg H, Sheppard D, Parton A, Armean IM, Trevanion SJ, Flicek P, Cunningham F. Ensembl variation resources. Database (Oxford) Published Online First: 2018. doi:10.1093/database/bay119

