## Supplementary Files for "GWAS of Chronic Dizziness in the Elderly Identifies Novel Loci Implicating *MLLT10, BPTF, LINC01225*, and *ROS1*"

#### SUPPLEMENTARY MATERIAL

##### Table of Contents

**eMethods**. Participants, phenotyping, assessment of balance and covariates in MVP, genotyping and imputation, relatedness, calculation of principal components (PCs), GWAS in MVP, estimating SNP-based heritability ($h^2_{SNP}$), colocalization of eQTL with GWAS results, replication in European meta-analysis from Iceland/Finland/UKB/US, CC-GWAS, fine-mapping, functional mapping and annotation, gene-based and gene set analysis with MAGMA, genetic correlations of tinnitus with other cochlear disorders, pheWAS of other disorders.

**Table e1.** Participants removed for in-patient and out-patient diagnoses.

**Table e2.** Linkage Disequilibrium Score on cochlear disorders and replication cohort.

**Table e3.** Case-case GWAS EU Balance compared to replication cohort.

**Table e4.** Gene-tissue expression in 53 tissues, including chromatin interactions.

**Table e5.** Functional analysis of lead SNPs.

**Table e6.** PheWAS results for lead SNPs.

**Figure e1.** Quantile-quantile plot of expected versus observed -log10 p-values for genome-wide association studies (GWAS) of chronic imbalance.

**Figure e2.** Regional association plots (Locus zoom) of top hit rs12779865 in trans-ancestry GWAS with annotations regarding pathogenicity, transcription-factor binding sites, and chromatin state.

**Figure e3.** Regional association plots (Locus zoom) of top hit rs72631329 in the trans-ancestry GWAS with annotations regarding pathogenicity, transcription-factor binding sites, and chromatin state.

**Figure e4.** Regional association plots (Locus zoom) of top hit rs10713223 in trans-ancestry GWAS with annotations regarding pathogenicity, transcription-factor binding sites, and chromatin state.

**Figure e5.** Regional association plots (Locus zoom) of top hit rs200689072 in trans-ancestry GWAS with annotations regarding pathogenicity, transcription-factor binding sites, and chromatin state.

**Figure e6.** Regional association plots (Locus zoom) of top hit rs10828248 in European (EU) GWAS with annotations regarding pathogenicity, transcription-factor binding sites, and chromatin state.

**Figure e7.** Regional association plots (Locus zoom) of top hit rs7221651 in EU GWAS with annotations regarding pathogenicity, transcription-factor binding sites, and chromatin state.

**Figure e8.** Regional association plots (Locus zoom) of top hit rs4791046 in EU >= age 50 GWAS with annotations regarding pathogenicity, transcription-factor binding sites, and chromatin state.

**Figure e9.** Regional association plots (Locus zoom) of top hit rs71717606 in Hispanic (LA) GWAS with annotations regarding pathogenicity, transcription-factor binding sites, and chromatin state.

**Figure e10.** Fine-mapping plots.

**Figure e11.** PheWAS on lead SNPs with annotation of top hits.

**Supplementary Methods:**

***Participants***

The Million Veteran Program (MVP) cohort consists of 819,307 participants (version 21_1) recruited since 2011. After written informed consent, participants fill out two basic health question surveys and have bloods drawn for genotyping. Information including ICD diagnostic codes, questionnaires, VA and active-duty health records are linked to de-identified ID#'s. Participants fill out basic and lifestyle questionnaires in regard to demographics, lifestyles, work, military experience, and self-reported health conditions. All participants provided written informed consent to participate.

***Phenotyping - Assessment of chronic imbalance and other variables in MVP***

"Minimal phenotyping" is defined as dependence on self-report, i.e., subjective "symptoms" rather than objective "signs" for case definitions and can degrade the quality and specificity of GWAS results.[19] Further, it has been shown that even studies that use a single diagnosis in the electronic health record can elicit a low sensitivity of 50% and specificity of 81%, leading to high rates of false positives in subsequent genome-wide association studies (GWAS).[19,20] We attempted to address this problem of self-report and individual examinations using multiple ICD codes, implying repeat examinations by a healthcare provider and symptoms of imbalance over a long period of time, i.e., six months or more.[21]

In an attempt to narrow the physiologic definition, the phenotyping was designed to remove acute vestibular diseases, as well as primary cerebellar and other intracranial diseases of imbalance. Although a large portion of chronic dizziness in the elderly includes the diagnosis of benign paroxysmal positional vertigo (BPPV),[15,16] this was excluded, as the primary focus was the aging cochlea. Other disqualifying diagnoses included acute and episodic vestibular syndromes such as Meniere's Disease, vertigo associated with migraine, cerebellar ataxias including Parkinson's Disease, and history of traumatic brain injury (TBI).

Our phenotype consisted of two diagnoses of dizziness, imbalance, or vertigo, at least 180 days apart. ICD's included were identified within the Veteran's Administration Informatics and Computing Infrastructure (VINCI) space via SQL tables of diagnoses and descriptions.

Excluded ICD's, both ICD9 and ICD10 were the following: 332.1, 334.x ataxias, 342.x hemiplegias, 386 Meniere's disease, 438.84 ataxia 780.2 syncope, 966.4 poisoning, E855.0 accidental overdose, E936.4 Parkinson's medications, G20 Parkinson's Disease, G21.x Secondary Parkinson's, G32.81 cerebellar ataxia, G60.2 neuropathy associated with hereditary ataxia, G80.x cerebral palsy and paralytic syndromes, G90.01 familial dysautonomia, G81.x hemiplegia, H81.01 Meniere's disease, H82.1 benign positional paroxysmal vertigo, H81.2 vestibular neuronitis, I69.x ataxia following subarachnoid hemorrhage, R27.0 ataxia, R55 syncope and collapse, T42.8x1A poisoning by Parkinson's medication, T67.1xxA heat syncope, T75.22xA external causes of spasticity, and R55 syncope. See Table e1.

Since a traumatic brain injury (TBI) can lead to vertigo, if the subject had a positive ICD for dizziness or vertigo AND had evidence of TBI, they were excluded. These diagnoses included S06.2x0A traumatic brain injury, S06.2x diffuse brain injury, S06.x intracranial injury, and V15.52 history of TBI.

Controls were those no evidence of imbalance in the electronic health record (EHR).

Here is a list of exclusions (note some participants fit into several categories):

- East and West Asians: 6,326
- < 180 days between diagnoses: 121,745
- Diagnoses:
  A. Acute diagnoses, such as Meniere's Disease, benign paroxysmal positional vertigo, vestibular neuronitis, viral labyrinthitis – 26,353
  B. Ataxias, including Parkinson's, cerebral ataxia, hereditary ataxias, following arachnoid hemorrhage – 27,768
  C. Cerebral palsy – 206
  D. Hemiplegia, cerebrovascular accident, stroke – 35,551
  E. Poisoning and overdose – 38,005
  F. Syncope, including heat syncope – 115,709
  G. Traumatic brain injury, intracranial injury, history of TBI – 54,722

***Genotyping and Imputation***

Analyses were based on the version 3 release of the MVP unimputed genetic dataset. Details of genotyping, quality control, and imputation of the MVP have been previously reported.[19] In brief, blood samples were gathered from subjects after written consent had been obtained, beginning in 2011. The Advanced Marker QC Pipeline excluded probeset calls from batches that fail advanced QC tests, as described previously (Hunter-Zinck, et al, 2020). Samples are stored at MAVERIC, VA Boston, MA. DNA was extracted and sent for genotyping at the Affymetrix Research Services Laboratory. Subjects were genotyped on either the Affymetrix Axiom or UK BiLEVE Axiom arrays, with approximately 4,700 samples per batch. Genotype calling was performed using a custom pipeline designed for biobank scale data. Routine quality control procedures were performed at each step of genotype extraction and calling. Quality control of marker genotypes included tests for batch and plate effects, deviations from Hardy-Weinberg equilibrium based on exact tests, sex effects, array effects, and discordance among technical replicates. Based on markers passing QC, subjects were removed for >2% missing genotype rate, discrepancy between self-reported sex and genetically determined sex, or excessive heterozygosity. Phasing was performed using SHAPEIT3 in partially overlapping chunks of 15,000 markers, with the 1000 Genomes Phase 3 dataset used as a reference panel. Chunks were merged using hapfuse. Data was imputed using IMPUTE4, using the combination of the 1000 Genomes Phase 3, UK10K, and Haplotype Reference Consortium panels, where the latter was preferentially used as the imputation reference.

***Relatedness***

Although REGENIE compensates for relatedness, nevertheless, related individuals were removed. Relatedness was estimated using KING. For analyses of unrelated subjects, from each pair with an estimated kinship coefficient > 0.0442 (3$^{rd}$ degree or closer), the individual with higher dizziness symptoms was retained, and for pairs with identical symptoms, one was chosen at random.

***Calculation of principal components (PCs) to address population stratification***

SNPs were excluded that had a MAF < 5%, HWE p > $1x10^{-3}$, call rate < 98%, were ambiguous (A/T, G/C), located in the MHC region (chr6, 25-35 MB) or chromosome 8 inversion (chr8, 7-13 MB). SNPs were pairwise LD-pruned ($r^2 > 0.2$) and a random set of ~100K markers was used for each subset to calculate PCs based on the smartPCA algorithm in EIGENSTRAT.

***GWAS in MVP***

REGENIE was run as described (Mbatchou et al., 2021). REGENIE is a machine-learning method using a mixed-model-based approach that uses ridge regression to combine a set of correlated predictors based on unimputed SNPs to fit a model calculated in Step 1. Since this phenotype is a binary trait, logistic regression with Firth correction was used. Covariates include 10 principal components, age, and sex.

REGENIE is a two-step machine learning method that accounts for relatedness. Step One consisted of analysis of MVP genotype array data only, where SNPs were partitioned into consecutive blocks, with ridge regression predictions generated from each block. Chunks consist of 219 MB. Predictions were then combined in a second ridge regression procedure and decomposed into separate chromosomes for leave-one-chromosome-out (LOCO) analysis. These LOCO predictions then became covariates in Step Two. In Step Two, Release 4 imputed data was used for cross validation. Firth logistic regression and saddle point approximation was used for this binary trait.

Summary statistics were filtered to MAF ≥ 1% and INFO score ≥ 0.6. Quantile-quantile (QQ) plots of expected versus observed $-\log_{10}$ *P*-values included genotyped and imputed SNPs at MAF ≥ 1%. The proportion of inflation of test statistics due to the actual polygenic signal (rather than other causes such as population stratification) was estimated as 1 - (1 - LDSC intercept - 1)/(mean observed chi-square - 1), using LD-score regression.[8] For primary analyses, genome-wide significance was declared at $P < 5 \times 10^{-8}$. Regional association plots were generated using LocusZoom with 400KB windows around the index variant with LD patterns calculated based on 1KGP EUR, AMR, or multiple reference populations for transancestry analysis.[9]

***Comparison to Iceland/Finland/UK/US cohort***

Methods of the Iceland/Finland/UK/US cohort GWAS are explained in detail in the original paper (REF 22). Briefly, the meta-analysis consisted of cohorts in Iceland (30,802 cases and 278,502 controls), the UK (9715 cases and 421,332 controls), the US (1888 cases and 24,961 controls), and Finland (5667 cases and 169,746 controls). The Ice/Fin/UKB/US phenotype had been defined as a single EHR diagnosis of vertigo, including ICD9 386.x and ICD10 of H81.xx., which includes acute syndromes.[22] GWAS summary stats from each cohort was then combined in a fixed-effects inverse variance model.

For GWAS, a logistic regression assuming an additive model was tested using deCODE. LD score regression to account for distribution inflation secondary to cryptic relatedness and population stratification. A fixed effect inverse variance method based on effect estimates and standard errors was used, where study groups were assumed to have a common odds ratio but allowed to have different population frequencies for alleles. Total number of variants were 62,056,310 spread over the four populations. GWAS significance was calculated using a weighted Bonferroni adjustment controlling for the family-wise error rate. In a random-effects method, a likelihood ratio test was performed on GWAS associations to test heterogeneity.

Variants had been mapped to NCBI Build38 in the Ice/Fin/UKB/US study, and for the current study, required lift-over to GrCh37.

***Estimating SNP-based heritability*** *($h^2_{SNP}$)*

SNP-based heritability estimates ($h^2_{SNP}$) were calculated using LDSC on GWAS summary data. Estimates were calculated for each version of the phenotype. Unconstrained regression intercepts were used to account for potential inclusion of related subjects and residual population stratification, and precomputed LD-scores from 1KGP EUR populations were used.

***Cohen's D***

Cohen's D was calculated to ascertain whether there was a difference in effect size between the European cohort and those over 50 years of age. Comparisons were computed for the lead SNP effect sizes in chromosomes 10 and 17, using beta for effect sizes, and converting standard error to standard deviation corrected for sample size in each cohort.

***CC-GWAS between Ice/Fin/UKB/US cohorts***

Case-case GWAS was performed to test for differences between SNPs of disorders. CC-GWAS is a method to test for genetic differences between allele frequencies of two disorders. Input consists of effect sizes, effective N's for cases and controls, P-values, population statistics and heritability for the disorders, and overlap between the two cohorts. For significant SNPS (P-value = 5.0e-10), output included statistically significant effect sizes reflecting direction and magnitude of effect, discards false positives, and measures genetic distance between the two disorders (Peyrot, 2021). Input consisted of effect sizes, effective N's for cases and controls, P-values, population statistics, heritability for the disorders, and overlap between the two cohorts.

***Fine-mapping with sum of single effects model.***

susieR v0.12.27 is a Bayesian method to assess uncertainty in which variables are selected, involving highly correlated variables. Originally formulated for linear regression, susieR has been verified for logistic regression. susieR provides "Credible Sets" of variables which include relevant SNPs and a posterior inclusion probability (PIP) for each variable based on effect sizes and linkage disequilibrium (LD) between SNPs. Minimum LD is 0.5 within each CS, and indicates that within each CS, one of the variables has a non-zero effect coefficient. An interval of 2.5 million base-pairs surrounding each significant locus was identified. This range was then downloaded from EnSembl in Data Slicer using these chromosome positions in GrCh38 and ancestries to match our lead SNPs from the 1000 genome. For example, since significant SNPs for the European cohort were identified in chromosomes 10 and 17, the ld matrix was fashioned from 1000 genomes of CEU individuals. Plink was then used to convert the .vcf file to plink binary, then linkage disequilibrium matrices were fashioned for use in susieR.

A fitted regression with summary statistics was fashioned for each significant locus, which created "credible sets" of potentially causal variants, based on posterior probability of 0.95, with 100 iterations and refinement of the model. Increasing the iterations did not improve the model.

***Functional mapping and annotation***

We used Functional Mapping and Annotation of genetic associations (FUMA) v1.3.5 (http://fuma.ctglab.nl/) to annotate GWAS data and obtain functional characterization of risk loci.[10] Annotations are based on human genome assembly GRCh37 (hg19). FUMA was used with default settings unless stated otherwise. The SNP2Gene module was used to define independent genomic risk loci and variants in LD with lead SNPs ($r^2 > 0.6$, calculated using ancestry appropriate 1KGP reference genotypes). SNPs in risk loci were mapped to protein coding genes with a 10kb window.

Functional consequences of SNPs were obtained by mapping the SNPs on their chromosomal position and reference alleles to databases containing known functional annotations, including ANNOVAR, Combined Annotation Dependent Depletion (CADD), RegulomeDB (RDB), and chromatin states in brain tissues/cell types. Next eQTL mapping was performed on significant (FDR < 0.05) SNP-gene pairs, mapping to GTEx v7 brain tissue, RNAseq data from the CommonMind Consortium and the BRAINEAC database. Chromatin interaction mapping was performed using built-in chromatin interaction data from the dorsolateral prefrontal cortex, hippocampus and neuronal progenitor cell line and adult and fetal cortex tissue. An FDR of $< 1x10^{-5}$ defined significant interactions, based on previous recommendations, modified to account for the differences in cell lines used here.

##### *Gene-based and gene set analysis with MAGMA*

Gene-based analysis was performed with the FUMA implementation of MAGMA.[11] SNPs were mapped to 18,222 protein coding genes. For each gene, its association with tinnitus was determined as the weighted mean squared test statistic of SNPs mapped to the gene, where LD patterns were calculated using ancestry appropriate 1KGP EUR reference genotypes. Significance of genes was set at a Bonferroni-corrected threshold of $P$ value = 0.05/18,222 = $2.7 \times 10^{-6}$.

To see if specific biological pathways were implicated in tinnitus, gene-based test statistics were used to perform a competitive set-based analysis of 10,894 pre-defined curated gene sets and GO terms obtained from MsigDB using MAGMA. Significance of pathways was set at a Bonferroni-corrected threshold of $P = 0.05/10,894 = 4.6 \times 10^{-6}$. To test if tissue-specific gene expression was associated with tinnitus, gene set-based analysis was also used with expression data from GTEx v7 RNA-seq and BrainSpan RNA-seq, where the expression of genes within specific tissues were used to define the gene properties used in the gene-set analysis model.

#### Table e1. Diagnoses Removed from Phenotype

| Description | In_patient | Out_patient | Total |
|---|---|---|---|
| Benign Paroxysmal Positional Vertigo | 3,506 | 40,126 | 41,233 |
| Meniere's Disease | 947 | 5,243 | 5,446 |
| Vestibular Neuronitis | 2,812 | 25,389 | 26,353 |
| Cerebral ataxia, Parkinson, hereditary ataxia | 8,521 | 25,185 | 27,768 |
| Cerebral Palsy | 45 | 174 | 219 |
| Hemiplegia, stroke | 16,461 | 28,412 | 35,551 |
| Poisoning and overdose | 16,838 | 28,367 | 38,005 |
| TBI, diffuse brain injury, intracranial injury | 12,007 | 50,775 | 54,722 |
| Syncope, including heat and orthostatic hypotension | 32,994 | 110,125 | 115,709 |
| TOTALS (may overlap) | 94,131 | 313,796 | 345,006 |

#### Table e2. Linkage disequilibrium scores on cochlear disorders and replication cohort.

| p1 | p2 | rg | se | Z-score | P value | h2_p1_liab | h2_p1_SE | h2_p2_liab | h2_p2_se | h2_int | h2_int_se | gcov_int | gcov_int_se |
|---|---|---|---|---|---|---|---|---|---|---|---|---|---|
| **balance** | **Skuladottir** | 0.6721 | 0.0734 | 9.1569 | 5.3433e-20 | 0.0469 | 0.0059 | 0.0666 | 0.0074 | 1.0053 | 0.0076 | 0.0051 | 0.0052 |
| **balance** | **tinnitus** | 0.2025 | 0.0833 | 2.4304 | 0.0151 | 0.0462 | 0.0066 | 0.1026 | 0.0066 | 1.0116 | 0.0074 | 0.0028 | 0.0059 |
| **balance** | **hearing** | 0.3148 | 0.0481 | 6.5495 | 5.7721e-11 | 0.0461 | 0.0059 | 0.1199 | 0.0006 | 1.0273 | 0.0094 | 0.0002 | 0.0053 |
| **tinnitus** | **hearing** | 0.5184 | 0.0309 | 16.7584 | 4.9136e-63 | 0.1071 | 0.0067 | 0.1193 | 0.0058 | 1.0282 | 0.0081 | 0.1544 | 0.0054 |
| **Skuladottir** | **tinnitus** | 0.1925 | 0.049 | 3.9242 | 8.7001e-05 | 0.067 | 0.0072 | 0.1066 | 0.0068 | 1.0098 | 0.0073 | 0.0283 | 0.0048 |
| **Skuladottir** | **hearing** | 0.1859 | 0.0491 | 3.7821 | 0.0002 | 0.067 | 0.0072 | 0.1208 | 0.006 | 1.0266 | 0.0087 | 0.0183 | 0.0052 |
| **balance** | **balance>49** | 0.9787 | 0.0063 | 154.2505 | 0.0 | 0.0459 | 0.0058 | 0.0482 | 0.007 | 1.0217 | 0.0075 | 0.9827 | 0.0074 |
| **EU Bal_90** | **EU Bal_180** | 1.0015 | 0.0035 | 288.075 | 0.0 | 0.0172 | 0.0024 | .0172 | .0024 | 1.0263 | 0.0100 | 0.9886 | 0.0098 |

**Abbreviations: pl, phenotype 1; p2, phenotype 2; rg, genomic correlation; se, standard error; h2, SNP inheritability on the liability scale; int, intercept, gcov, genetic covariance; EU Bal_90 European cohort for 2 diagnoses of dizziness greater than 90 days apart; EU Bal_180 European cohort for 2 diagnoses greater than 180 days apart.**

Balance study – Trans-ancestry GWAS from Million Veterans Program

Tinnitus study – European GWAS performed in UK Biobank (Clifford, et. al, 2020)

Hearing study – European GWAS performed in UK Biobank (Wells, et al, 2019)

Skuladottir, et al. – European meta-analysis GWAS on Iceland/Finland/UKBiobank/US for diagnosis of vertigo (Skuladottir, et al., 2021)

**Table e3. Case-case European Balance GWAS compared to Replication Cohort[1]**

| SNP | CHR | BP | EA | NEA | OLS_beta | OLS_se | OLS_pval | Exact_beta | Exact_se | Exact_pval | CCGWAS_signif |
|---|---|---|---|---|---|---|---|---|---|---|---|
| rs403983 | 19 | 23484776 | G | A | -2.97E-03 | 5.27E-04 | 1.81E-08 | -1.82E-02 | 3.22E-03 | 1.55E-08 | 1 |
| rs17473980 | 19 | 23512665 | G | T | -5.24E-03 | 5.27E-04 | 2.82E-23 | -3.22E-02 | 3.22E-03 | 1.72E-23 | 1 |
| rs11667562 | 19 | 23513267 | T | G | -4.98E-03 | 5.27E-04 | 3.80E-21 | -3.06E-02 | 3.22E-03 | 2.55E-21 | 1 |
| rs12151189 | 19 | 23517735 | C | T | -4.97E-03 | 5.27E-04 | 4.13E-21 | -3.05E-02 | 3.22E-03 | 2.78E-21 | 1 |
| rs35367868 | 19 | 23527117 | C | T | -4.99E-03 | 5.27E-04 | 2.90E-21 | -3.07E-02 | 3.22E-03 | 1.94E-21 | 1 |
| rs295390 | 19 | 23528350 | T | C | -4.91E-03 | 5.27E-04 | 1.31E-20 | -3.02E-02 | 3.22E-03 | 8.40E-21 | 1 |
| rs1020075 | 19 | 23540217 | T | C | -4.97E-03 | 5.27E-04 | 4.09E-21 | -3.05E-02 | 3.22E-03 | 2.75E-21 | 1 |
| rs35804934 | 19 | 23557070 | A | G | -3.24E-03 | 5.27E-04 | 8.51E-10 | -1.98E-02 | 3.22E-03 | 8.67E-10 | 1 |
| rs641702 | 19 | 23593010 | G | T | -5.11E-03 | 5.27E-04 | 3.38E-22 | -3.14E-02 | 3.22E-03 | 2.23E-22 | 1 |
| rs147995216 | 19 | 23601870 | G | A | -5.08E-03 | 5.27E-04 | 5.49E-22 | -3.12E-02 | 3.22E-03 | 3.61E-22 | 1 |
| rs12983355 | 19 | 23607974 | G | T | -4.40E-03 | 5.27E-04 | 6.75E-17 | -2.70E-02 | 3.22E-03 | 5.05E-17 | 1 |
| rs12985969 | 19 | 23611666 | A | C | -5.08E-03 | 5.27E-04 | 6.04E-22 | -3.12E-02 | 3.22E-03 | 4.00E-22 | 1 |
| rs2219838 | 19 | 23634712 | C | T | -3.05E-03 | 5.27E-04 | 7.62E-09 | -1.86E-02 | 3.22E-03 | 7.63E-09 | 1 |
| rs10413718 | 19 | 23655398 | T | G | -4.06E-03 | 5.27E-04 | 1.34E-14 | -2.49E-02 | 3.22E-03 | 1.02E-14 | 1 |
| rs8108305 | 19 | 23665224 | T | C | -4.06E-03 | 5.27E-04 | 1.39E-14 | -2.49E-02 | 3.22E-03 | 1.06E-14 | 1 |
| rs10420240 | 19 | 23670982 | G | A | -3.95E-03 | 5.27E-04 | 6.40E-14 | -2.43E-02 | 3.22E-03 | 5.03E-14 | 1 |

**Abbr: SNP, single nucleotide polymorphism; CHR chromosome; EA, effect allele; NEA, non-effect allele; OLS ordinary least-squares; pval, P-value; se, standard error; CCGWAS signif, case-case genome-wide association study – a comparison of alleles at each SNP.**

**[1] Based on comparison of 7,600,837 overlapping SNPs between EU GWAS and Ice/UK/US/Fin replication.**

**Table e4. Gene-Tissue Expression in 53 tissues with addition of chromatin interactions.**

| VARIABLE | BETA | BETA_STD | SE | P |
|---|---|---|---|---|
| Brain_Cerebellar_Hemisphere | 0.024937 | 0.050001 | 0.0062109 | 2.99E-05 |
| Brain_Cerebellum | 0.024979 | 0.049545 | 0.0063991 | 4.76E-05 |
| Brain_Frontal_Cortex_BA9 | 0.019148 | 0.035243 | 0.0070027 | 0.0031278 |
| Brain_Cortex | 0.019399 | 0.03534 | 0.0072543 | 0.0037498 |
| Pituitary | 0.021851 | 0.040672 | 0.0091688 | 0.0085881 |
| Brain_Anterior_cingulate_cor... | 0.015181 | 0.026789 | 0.0073387 | 0.019299 |
| Brain_Spinal_cord_cervical_c... | 0.016329 | 0.029318 | 0.0088446 | 0.032442 |
| Brain_Putamen_basal_ganglia | 0.013628 | 0.023169 | 0.0080772 | 0.045788 |
| Brain_Hippocampus | 0.01341 | 0.022686 | 0.0081126 | 0.049181 |
| Brain_Caudate_basal_ganglia | 0.012384 | 0.021369 | 0.0080142 | 0.061152 |
| Brain_Amygdala | 0.010826 | 0.018424 | 0.0079675 | 0.087128 |
| Brain_Nucleus_accumbens_basa... | 0.01035 | 0.017938 | 0.0077289 | 0.090277 |
| Brain_Hypothalamus | 0.0095061 | 0.016342 | 0.0082465 | 0.12452 |
| Brain_Substantia_nigra | 0.0098406 | 0.016924 | 0.0087676 | 0.13086 |
| Prostate | 0.008112 | 0.015366 | 0.012212 | 0.25326 |
| Whole_Blood | 0.0029887 | 0.0054018 | 0.0057267 | 0.30088 |
| Liver | 0.0032138 | 0.0057853 | 0.0065339 | 0.31141 |
| Small_Intestine_Terminal_Ile... | 0.0048361 | 0.0088795 | 0.0098472 | 0.31168 |
| Stomach | 0.0058953 | 0.010698 | 0.012355 | 0.31663 |
| Muscle_Skeletal | 0.0030005 | 0.005676 | 0.0071178 | 0.33668 |
| Ovary | 0.00029856 | 0.00060469 | 0.0099363 | 0.48801 |
| Heart_Left_Ventricle | -0.00099425 | -0.0016592 | 0.0093169 | 0.54249 |
| Bladder | -0.0031143 | -0.0060624 | 0.013287 | 0.59266 |
| Heart_Atrial_Appendage | -0.0030471 | -0.0054671 | 0.0097693 | 0.62244 |
| Pancreas | -0.002821 | -0.004716 | 0.0086482 | 0.62786 |
| Adrenal_Gland | -0.0038153 | -0.007359 | 0.010246 | 0.64519 |
| Cervix_Endocervix | -0.0054448 | -0.010763 | 0.011856 | 0.67697 |
| Cells_EBV-transformed_lympho... | -0.0023749 | -0.0051821 | 0.0050178 | 0.682 |

| | | | | |
|---|---|---|---|---|
| Colon_Transverse | -0.0076424 | -0.014033 | 0.011872 | 0.74012 |
| Testis | -0.0037007 | -0.006376 | 0.0056747 | 0.74284 |
| Esophagus_Gastroesophageal_J... | -0.010153 | -0.019969 | 0.013208 | 0.77896 |
| Uterus | -0.0093772 | -0.019085 | 0.011163 | 0.79954 |
| Spleen | -0.0072687 | -0.014393 | 0.0074528 | 0.83528 |
| Cervix_Ectocervix | -0.01484 | -0.02905 | 0.012377 | 0.88473 |
| Lung | -0.011574 | -0.022345 | 0.009339 | 0.89239 |
| Esophagus_Muscularis | -0.015937 | -0.031409 | 0.012793 | 0.89356 |
| Artery_Tibial | -0.012876 | -0.026489 | 0.010323 | 0.89385 |
| Kidney_Medulla | -0.011939 | -0.022052 | 0.0095334 | 0.89476 |
| Fallopian_Tube | -0.015477 | -0.029978 | 0.012199 | 0.89772 |
| Colon_Sigmoid | -0.017027 | -0.033275 | 0.012915 | 0.9063 |
| Kidney_Cortex | -0.013487 | -0.023676 | 0.0092842 | 0.92684 |
| Skin_Sun_Exposed_Lower_leg | -0.012154 | -0.023554 | 0.0078837 | 0.9384 |
| Artery_Aorta | -0.016264 | -0.033163 | 0.010519 | 0.93895 |
| Skin_Not_Sun_Exposed_Suprapu... | -0.012257 | -0.023611 | 0.0078599 | 0.94055 |
| Minor_Salivary_Gland | -0.016657 | -0.030963 | 0.009838 | 0.95478 |
| Thyroid | -0.017333 | -0.034282 | 0.0098778 | 0.96034 |
| Nerve_Tibial | -0.01882 | -0.037828 | 0.010344 | 0.96556 |
| Cells_Cultured_fibroblasts | -0.014839 | -0.03178 | 0.0066835 | 0.98679 |
| Adipose_Subcutaneous | -0.024551 | -0.048951 | 0.011017 | 0.98707 |
| Vagina | -0.025374 | -0.048454 | 0.011223 | 0.98811 |
| Breast_Mammary_Tissue | -0.030297 | -0.058188 | 0.012921 | 0.99048 |
| Esophagus_Mucosa | -0.018348 | -0.035437 | 0.0077716 | 0.99088 |
| Adipose_Visceral_Omentum | -0.028133 | -0.054587 | 0.011358 | 0.99337 |
| Artery_Coronary | -0.030466 | -0.060584 | 0.012208 | 0.99371 |

**European GWAS in MVP. Significant p-value = 0.05/53 = 9.4 x $10^{-04}$. MEAN_SAMPLE_SIZE = 54557. TOTAL_GENES = 17236**

#### Table e5. Functional analysis of lead SNPs

| Ancestry | Lead Variant | P-value | Risk Locus | # SNPS in LD > .6 | Predicted Genes in Locus | SNPS in LD[a] CADD[b] > 12.37 |
|---|---|---|---|---|---|---|
| Trans-ancestry | rs12779865 | 3.65E-08 | 10:21769994-22285371 | 91 | SKIDA1, MLLT10, DNAJC1, CASC10 | rs12770228 |
| | rs72631329 | 7.61E-09 | 17:65814382-66098154 | 333 | BPTF, C17orf58, KPNA2 | rs12452511, rs1976053, rs75360723, rs75360723, rs4318247, + 5 more |
| | rs10713223 | 3.11E-08 | 19:23490203-23642411 | 89 | ZNF91 | rs428549, rs397756857, rs10585848 |
| | rs200689072 | 2.43E-08 | 21:18512554-18548915 | 16 | none | rs8134340 |
| CEU | rs10828248 | 3.17E-08 | 10:21766969-22288132 | 145 | SKIDA1; MLLT10; DNAJC1; CASC10 | rs12256551, rs946711, rs12770228, rs66955492, rs11012786 |
| | rs7221651 | 3.71E-10 | 17:65764164-66098154 | 404 | BPTF, C17orf58, KPNA2 | rs12452511, rs1976053, rs75360723, rs11656743, rs75868869, + 3 more |
| CEU > 50 | rs10828248 | 1.49E-08 | 10:21766969-22288132 | 145 | SKIDA1, MLLT10, DNAJC1, CASC10 | rs12452511, rs1976053, rs75360723, rs75360723, rs4318247, + 5 more |
| | rs4791046 | 5.24E-09 | 17:65764164-66096529 | 351 | BPTF, C17orf58, KPNA2 | rs12452511, rs1976053, rs75360723, rs11656743, rs75868869, + 3 more |
| MXL | rs71717606 | 4.84E-10 | 6:117683821-117822159 | 169 | GOPC, ROS1, DCBLD1 | rs2229079, rs368013630, rs72951624, rs9489171, rs17079187, rs56115066 |

Abbreviations: SNPs, single nucleotide polymorphisms; LD linkage disequilibrium;, eQTL, expression quantitative trait locus; Hi-C, hybridization capture of chromatin interactions.

[a] When > 5 are in this category, only the top 5 are listed.

[b] CADD is an *in silico* prediction of variant pathogenicity. >12.37 is a suggested threshold for pathogenicity.

[c] RegulomeDBscore < 5 indicates increased possibility of regulatory element binding sites.

[d] Hi-C provides evidence for chromatin interaction.

#### Table e5. Functional analysis of lead SNPs (continued)

| Ancestry | Lead Variant | RegulomeDB[c] < 5 |
|---|---|---|
| Trans | rs12779865 | rs12770228, rs12357321, rs11012726, rs11012732, rs10828252, … + 5 |
| | rs72631329 | rs3936134, rs12451721, rs7214119, rs62084208, rs115210018, + 27 more |
| | rs10713223 | rs11882589, rs641702, rs603909, rs695017, rs586413, + 3 more |
| | rs200689072 | none |
| CEU | rs10828248 | rs946711, rs10828249, rs10828247, rs2807986, rs12251016, + 19 more |
| | rs7221651 | rs3936134, rs12451721, rs7214119, rs62084208, rs12452511, + 36 more |
| CEU > 50 | rs10828248 | rs946711, rs10828249, rs10828247, rs2807986, rs12251016, + 19 more |
| | rs4791046 | rs3936134, rs12451721, rs7214119, rs62084208, rs62084208, + 29 more |
| MXL | rs71717606 | rs72965306, rs72965317, rs72969456, rs72959668, rs72959686, rs56741692, rs55977400, rs59267160, rs56115066 |

**Table e5 (continued)**

| Ancestry | Lead Variant | eQTL (13 neural tissue/cell lines) GTEx/v8 | Hi-C[d] in tissue cell line datasets |
|---|---|---|---|
| Trans | rs12779865 | CASC1 in cerebellum | SKIDA1 intrachromosomal adult cortex, COMMD3-BMI1 in hippocampus and dorsolateral prefrontal cortex, SPAG6 intrachromosomal in hippocampus, and DPFC |
| | rs72631329 | KPNA2 Cerebellar hemisphere, amygdala, nucleus accumbens basal gangia; BPTF cerebellum, cerebellar hemisphere | |
| | rs10713223 | none | none |
| | rs200689072 | none | none |
| CEU | rs10828248 | MLLT10 anterior cinguate cortex, CASC10 cerebellum | MLLT10 hippocampus, DNAJC1 dorsolateral prefrontal cortex, hippocampus |
| | rs7221651 | KPNA2 caudate basal ganglia, amygdala, nucleus aNoneccumbens basal ganglia, BPTF cerebellum, cerebellar hemisphere | none |
| CEU > 50 | rs10828248 | MLLT10 anterior cinguate cortex, CASC10 cerebellum | MLLT10 hippocampus, DNAJC1 dorsolateral prefrontal cortex, hippocampus |
| | rs4791046 | KPNA2 in anterior cingulate cortex, brain cortex; BPTF cerebellum, cerebellar hemisphere | |
| MXL | rs71717606 | ROS1, GOPC, DCBLD1 in cortex, GOPC, DCBLD1 in frontal cortex | DCBLD1 intrachromosomal in hippocampus |

### Table e6. PheWAS results for lead SNPs.

#### Chr 6 rs72965321 Significant P-value 4.31e-04

| PMID | Year | Domain | Trait | P-value | N | EA | NEA |
|---|---|---|---|---|---|---|---|
| 31427789 | 2019 | Dermatological | Skin colour | 1.87E-05 | 381433 | C | T |
| 31427789 | 2019 | Nutritional | Coffee intake | 5.12E-05 | 358093 | C | T |
| https://doi.org/10.1101/288626 | 2019 | Neurological | Cingulum (hippocampus) mode of anisotropy | 8.44E-05 | 17706 | T | C |
| 30573740 | 2018 | Dermatological | Male pattern baldness (BOLT LMM infinitesimal mixed model) | 2.30E-04 | 205327 | T | C |

#### Chr 10 rs7894565 Significant P-value 2.94E-04

| PMID | Year | Domain | Trait | P-value | N | EA | NEA |
|---|---|---|---|---|---|---|---|
| doi.org/10.1101/288563 | 2019 | Neurological | Fornix (column and body of fornix) fractiona anisotropy | 3.39E-11 | 17706 | C | T |
| doi.org/10.1101/288651 | 2019 | Neurological | Fornix (column and body of fornix) radial diusivities | 1.73E-09 | 17706 | T | C |
| doi.org/10.1101/288607 | 2019 | Neurological | Fornix (column and body of fornix) mean diusivities | 2.64E-09 | 17706 | T | C |
| doi.org/10.1101/288590 | 2019 | Neurological | Posterior limb of internal capsule axial diusivities | 4.87E-09 | 17706 | T | C |
| 30664634 | 2019 | Metabolic | Arms-arm fat ratio (female) | 6.54E-09 | 195068 | C | T |
| doi.org/10.1101/288638 | 2019 | Neurological | Superior corona radiata mode of anisotropy | 8.22E-09 | 17706 | T | C |
| doi.org/10.1101/288585 | 2019 | Neurological | Fornix (column and body of fornix) axial diusivities | 2.72E-08 | 17706 | T | C |
| 30531941 | 2018 | Activities | Overall activity | 7.30E-08 | 91105 | T | C |
| doi.org/10.1101/288634 | 2019 | Neurological | Posterior limb of internal capsule mode of anisotropy | 1.26E-07 | 17706 | T | C |
| 27863252 | 2016 | Immunological | Monocyte count (three-way meta) | 3.66E-07 | 170721 | C | T |
| 27863252 | 2016 | Immunological | Monocyte percentage of white cells (three-way meta) | 4.41E-07 | 170494 | C | T |
| 30108127 | 2018 | Metabolic | Body Mass Index | 6.90E-07 | 334487 | C | T |
| 31676860 | 2019 | Neurological | Left accumbens area | 8.04E-07 | 19629 | C | T |
| 31676860 | 2019 | Neurological | Left accumbens area | 1.17E-06 | 21821 | C | T |
| 27863252 | 2016 | Immunological | Granulocyte percentage of myeloid white cells (three-way meta) | 1.68E-06 | 169545 | T | C |
| 31676860 | 2019 | Neurological | Total brain volume | 1.84E-06 | 21821 | T | C |
| 31676860 | 2019 | Neurological | Total brain volume | 2.10E-06 | 19629 | T | C |
| doi.org/10.1101/288572 | 2019 | Neurological | Superior corona radiata fractional anisotropy | 2.93E-06 | 17706 | T | C |
| 27863252 | 2016 | Immunological | Monocyte percentage of white cells (two-way meta) | 3.67E-06 | 131305 | C | T |
| doi.org/10.1101/288568 | 2019 | Neurological | Posterior limb of internal capsule fractional anisotropy | 4.15E-06 | 17706 | T | C |

| | | | | | | | |
|---|---|---|---|---|---|---|---|
| doi.org/10.1101/288633 | 2019 | Neurological | Posterior corona radiata mode of anisotropy | 4.37E-06 | 17706 | T | C |
| 27863252 | 2016 | Immunological | Monocyte count (two-way meta) | 5.17E-06 | 131544 | C | T |
| doi.org/10.1101/288594 | 2019 | Neurological | Superior corona radiata axial diusivities | 6.06E-06 | 17706 | T | C |
| 27863252 | 2016 | Immunological | Granulocyte percentage of myeloid white cells (two-way meta) | 1.24E-05 | 130543 | T | C |
| doi.org/10.1101/288622 | 2019 | Neurological | Anterior limb of internal capsule mode of anisotropy | 1.36E-05 | 17706 | T | C |
| 31676860 | 2019 | Neurological | 3rd ventricle | 1.53E-05 | 19629 | T | C |
| 27046643 | 2016 | Environment | Educational attainment | 1.79E-05 | 111114 | T | C |
| doi.org/10.1101/288629 | 2019 | Neurological | Fornix (column and body of fornix) mode of anisotropy | 1.80E-05 | 17706 | C | T |
| 31676860 | 2019 | Neurological | Left lateral ventricle | 2.05E-05 | 19629 | T | C |
| 31676860 | 2019 | Neurological | 3rd ventricle | 2.80E-05 | 21821 | T | C |

**Chr17 Significant P-value 8.98E-05 rs7221651**

| PMID | Year | Domain | Trait | P-value | N | EA | NEA |
|---|---|---|---|---|---|---|---|
| 31427789 | 2019 | Metabolic | Impedance measures - Trunk fat mass | 7.05E-30 | 379578 | C | T |
| 31427789 | 2019 | Metabolic | Impedance measures - Trunk fat percentage | 4.63E-27 | 379600 | C | T |
| 31427789 | 2019 | Metabolic | Impedance measures - Whole body fat mass | 8.14E-26 | 379203 | C | T |
| 31427789 | 2019 | Metabolic | Impedance measures - Body fat percentage | 1.23E-25 | 379615 | C | T |
| 31427789 | 2019 | Metabolic | Weight | 5.56E-23 | 385473 | C | T |
| 31427789 | 2019 | Metabolic | Impedance measures - Weight | 7.11E-23 | 379840 | C | T |
| 31427789 | 2019 | Metabolic | Waist circumference | 9.77E-22 | 385932 | C | T |
| 31427789 | 2019 | Metabolic | Impedance measures - Arm fat percentage (left) | 9.01E-20 | 379699 | C | T |
| 31427789 | 2019 | Metabolic | Impedance measures - Leg fat mass (right) | 1.23E-19 | 379802 | C | T |
| 31427789 | 2019 | Metabolic | Impedance measures - Leg fat mass (left) | 1.44E-19 | 379783 | C | T |
| 31427789 | 2019 | Metabolic | Impedance measures - Leg fat percentage (left) | 1.45E-19 | 379786 | C | T |
| 31427789 | 2019 | Metabolic | Impedance measures - Leg fat percentage (right) | 1.27E-18 | 379806 | C | T |
| 31427789 | 2019 | Metabolic | Impedance measures - Arm fat percentage (right) | 1.41E-18 | 379752 | C | T |
| 31427789 | 2019 | Metabolic | Impedance measures - Arm fat mass (right) | 2.55E-18 | 379725 | C | T |
| 31427789 | 2019 | Metabolic | Impedance measures - Arm fat mass (left) | 1.82E-17 | 379663 | C | T |
| 27863252 | 2016 | Immunological | Eosinophil count (three-way meta) | 2.22E-17 | 172275 | C | T |
| 31427789 | 2019 | Metabolic | Hip circumference | 9.33E-17 | 385887 | C | T |
| 27863252 | 2016 | Immunological | Eosinophil percentage of white cells (three-way meta) | 1.66E-16 | 172378 | C | T |
| 27863252 | 2016 | Immunological | Sum eosinophil basophil count (three-way meta) | 4.05E-16 | 171771 | C | T |
| 27863252 | 2016 | Immunological | Eosinophil count (two-way meta) | 1.70E-15 | 131999 | C | T |
| 27863252 | 2016 | Immunological | Eosinophil percentage of granulocytes (three-way meta) | 1.77E-15 | 170536 | C | T |
| 27863252 | 2016 | Immunological | Eosinophil percentage of white cells (two-way meta) | 1.77E-15 | 132052 | C | T |
| 27863252 | 2016 | Immunological | Sum eosinophil basophil count (two-way meta) | 2.12E-14 | 131557 | C | T |
| 27863252 | 2016 | Immunological | Eosinophil percentage of granulocytes (two-way meta) | 3.08E-14 | 131525 | C | T |
| 31427789 | 2019 | Metabolic | Impedance measures - Basal metabolic rate | 4.54E-14 | 379821 | C | T |
| 27863252 | 2016 | Immunological | Neutrophil percentage of granulocytes (three-way meta) | 8.86E-14 | 170672 | T | C |
| 31427789 | 2019 | Metabolic | Impedance measures - Leg fat-free mass (left) | 1.30E-13 | 379766 | C | T |
| 31427789 | 2019 | Metabolic | Impedance measures - Leg predicted mass (left) | 1.70E-13 | 379761 | C | T |
| 31427789 | 2019 | Metabolic | Impedance measures - Body Mass Index (BMI) | 2.61E-13 | 379831 | C | T |
| 30239722 | 2018 | Metabolic | Body Mass Index | 5.19E-13 | 806834 | C | T |
| 31427789 | 2019 | Metabolic | Body Mass Index | 5.46E-13 | 385336 | C | T |
| 30664634 | 2019 | Metabolic | Legs-leg fat ratio (female) | 6.07E-13 | 195068 | T | C |
| 27863252 | 2016 | Immunological | Neutrophil percentage of granulocytes (two-way meta) | 9.21E-13 | 131660 | T | C |
| 31427789 | 2019 | Skeletal | Standing height | 1.16E-12 | 385748 | C | T |
| 31427789 | 2019 | Metabolic | Impedance measures - Leg predicted mass (right) | 2.65E-12 | 379793 | C | T |
| 31427789 | 2019 | Metabolic | Impedance measures - Leg fat-free mass (right) | 2.78E-12 | 379793 | C | T |
| 30239722 | 2018 | Metabolic | Waist-hip ratio | 4.06E-12 | 697734 | C | T |

| | | | | | | | |
|---|---|---|---|---|---|---|---|
| doi.org/10.1101/261081 | 2018 | Reproduction | Number of sexual partners | 1.14E-11 | 370711 | C | T |
| 31427789 | 2019 | Metabolic | Impedance measures - Arm predicted mass (right) | 2.43E-11 | 379716 | C | T |
| 31427789 | 2019 | Metabolic | Impedance measures - Arm fat-free mass (right) | 3.33E-11 | 379723 | C | T |
| 31427789 | 2019 | Metabolic | Impedance measures - Whole body water mass | 3.49E-11 | 379835 | C | T |
| 31427789 | 2019 | Metabolic | Impedance measures - Whole body fat-free mass | 4.56E-11 | 379804 | C | T |
| 30297969 | 2018 | Endocrine | Type 2 Diabetes | 1.50E-10 | 898130 | C | T |
| 31427789 | 2019 | Metabolic | Impedance measures - Arm predicted mass (left) | 2.78E-10 | 379638 | C | T |
| 31427789 | 2019 | Metabolic | Impedance measures - Arm fat-free mass (left) | 5.54E-10 | 379653 | C | T |
| 29970889 | 2018 | Psychiatric | Loneliness (MTAG) | 6.20E-09 | 487647 | C | T |
| 31427789 | 2019 | Nutritional | Major dietary changes in the last 5 years | 8.65E-09 | 385587 | C | T |
| 30239722 | 2018 | Metabolic | Body Mass Index (female) | 2.52E-08 | 434794 | C | T |
| 30664634 | 2019 | Metabolic | Trunk-trunk fat ratio (female) | 2.67E-08 | 195068 | C | T |
| 30804560 | 2019 | Respiratory | FEV1/FVC ratio | 2.71E-08 | 400102 | C | T |
| 30239722 | 2018 | Metabolic | Waist-hip ratio (female) | 3.00E-08 | 381152 | C | T |
| 28892062 | 2017 | Metabolic | Body Mass Index | 3.07E-08 | 158284 | T | C |
| 31427789 | 2019 | Metabolic | Impedance measures - Trunk fat-free mass | 3.40E-08 | 379507 | C | T |
| 30804560 | 2019 | Respiratory | FEV1/FVC ratio | 4.10E-08 | 321047 | C | T |
| 29970889 | 2018 | Psychiatric | Loneliness | 5.30E-08 | 445024 | C | T |
| 31427789 | 2019 | Metabolic | Impedance measures - Trunk predicted mass | 7.32E-08 | 379469 | C | T |
| doi.org/10.1101/288628 | 2019 | Neurological | External capsule mode of anisotropy | 7.95E-08 | 17706 | C | T |
| doi.org/10.1101/288632 | 2019 | Neurological | Inferior fronto-occipital fasciculus mode of anisotropy | 1.22E-07 | 17706 | C | T |
| 29403010 | 2018 | Metabolic | Alanine aminotransferase | 1.97E-07 | 134182 | C | T |
| doi.org/10.1101/288618 | 2019 | Neurological | Superior longitudinal fasciculus mean diusivities | 3.26E-07 | 17706 | C | T |
| doi.org/10.1101/288611 | 2019 | Neurological | Posterior corona radiata mean diusivities | 7.04E-07 | 17706 | C | T |
| 31427789 | 2019 | Skeletal | Comparative height size at age 10 | 8.22E-07 | 380167 | C | T |
| doi.org/10.1101/288662 | 2019 | Neurological | Superior longitudinal fasciculus radial diusivities | 8.63E-07 | 17706 | C | T |
| doi.org/10.1101/261081 | 2018 | Activities | First PC of the four riskly behaviours | 9.02E-07 | 315894 | C | T |
| 31427789 | 2019 | Activities | Overall health rating | 1.01E-06 | 384850 | T | C |
| 31427789 | 2019 | Nutritional | Never eat eggs, dairy, wheat, sugar: I eat all of the above | 1.22E-06 | 384986 | T | C |
| 31676860 | 2019 | Neurological | Left caudal anterior cingulate | 1.24E-06 | 21821 | T | C |
| 31676860 | 2019 | Neurological | Left caudal anterior cingulate | 1.97E-06 | 19629 | T | C |
| doi.org/10.1101/288601 | 2019 | Neurological | Average across all tracts mean diusivities | 2.02E-06 | 17706 | C | T |
| 29403010 | 2018 | Metabolic | Triglyceride | 2.09E-06 | 105597 | C | T |
| 28892062 | 2017 | Metabolic | Body Mass Index (male) | 2.20E-06 | 85894 | T | C |
| 30239722 | 2018 | Metabolic | Body Mass Index (male) | 2.24E-06 | 374756 | C | T |

| | | | | | | | |
|---|---|---|---|---|---|---|---|
| 31427789 | 2019 | Social Interactions | Frequency of friend/family visits | 2.81E-06 | 383941 | T | C |
| doi.org/10.1101/288555 | 2019 | Neurological | Anterior corona radiata fractional anisotropy | 3.02E-06 | 17706 | T | C |
| doi.org/10.1101/288643 | 2019 | Neurological | Anterior corona radiata radial diusivities | 3.59E-06 | 17706 | C | T |
| 30108127 | 2018 | Metabolic | Body Mass Index | 3.70E-06 | 334487 | C | T |
| 31427789 | 2019 | Skeletal | Sitting height | 4.05E-06 | 385393 | C | T |
| 31427789 | 2019 | Psychiatric | Frequency of tiredness / lethargy in last 2 weeks | 4.26E-06 | 375053 | C | T |
| doi.org/10.1101/288645 | 2019 | Neurological | Average across all tracts radial diusivities | 4.49E-06 | 17706 | C | T |
| 30531953 | 2018 | Neurological | Epilepsy | 4.56E-06 | 44889 | C | T |
| doi.org/10.1101/288599 | 2019 | Neurological | Anterior corona radiata mean diusivities | 4.68E-06 | 17706 | C | T |
| 30598549 | 2018 | Body Structures | Fractures | 5.00E-06 | 426795 | C | T |
| 29403010 | 2018 | Metabolic | Gamma-glutamyl transferase | 6.04E-06 | 118309 | C | T |
| 29970889 | 2018 | Social Interactions | Social support - Leisure/social activities: Religious group | 6.60E-06 | 452302 | T | C |
| doi.org/10.1101/288574 | 2019 | Neurological | Superior longitudinal fasciculus fractional anisotropy | 9.11E-06 | 17706 | T | C |
| doi.org/10.1101/288655 | 2019 | Neurological | Posterior corona radiata radial diusivities | 9.88E-06 | 17706 | C | T |
| 31427789 | 2019 | Activities | Number of treatments/medications taken | 1.23E-05 | 386581 | C | T |
| 30054458 | 2018 | Endocrine | Type 2 Diabetes | 1.65E-05 | 659256 | C | T |
| 31427789 | 2019 | Reproduction | Age first had sexual intercourse | 1.90E-05 | 339614 | T | C |
| doi.org/10.1101/288584 | 2019 | Neurological | External capsule axial diusivities | 2.05E-05 | 17706 | C | T |
| doi.org/10.1101/288579 | 2019 | Neurological | Average across all tracts axial diusivities | 2.19E-05 | 17706 | C | T |
| 30048462 | 2018 | Skeletal | Heel bone mineral density | 2.50E-05 | 394929 | T | C |
| doi.org/10.1101/288658 | 2019 | Neurological | Retrolenticular part of internal capsule radial diusivities | 2.78E-05 | 17706 | C | T |
| 31427789 | 2019 | Psychiatric | Loneliness, isolation | 3.02E-05 | 380317 | C | T |
| 31427789 | 2019 | Activities | Health satisfaction | 3.07E-05 | 128745 | T | C |
| 31676860 | 2019 | Neurological | White matter | 3.54E-05 | 21821 | C | T |
| 31427789 | 2019 | Nutritional | Never eat eggs, dairy, wheat, sugar: Sugar or foods/drinks containing sugar | 3.62E-05 | 384986 | C | T |
| doi.org/10.1101/288557 | 2019 | Neurological | Average across all tracts fractional anisotropy | 3.91E-05 | 17706 | T | C |
| 29500382 | 2018 | Psychiatric | Loneliness, isolation (LONE) | 4.60E-05 | 267190 | C | T |
| 31427789 | 2019 | Activities | Types of physical activity in last 4 weeks: Light DIY (eg: pruning, watering the lawn) | 4.62E-05 | 384450 | T | C |
| /10.1101/288613 | 2019 | Neurological | Posterior thalamic radiation (include optic radiation) mean diusivities | 5.12E-05 | 17706 | C | T |
| 31427789 | 2019 | Body Structures | Fractured/broken bones in last 5 years | 5.56E-05 | 384446 | C | T |
| doi.org/10.1101/288657 | 2019 | Neurological | Posterior thalamic radiation (include optic radiation) radial diusivities | 5.84E-05 | 17706 | C | T |

| | | | | | | | |
|---|---|---|---|---|---|---|---|
| doi.org/10.11 01/288614 | 2019 | Neurological | Retrolenticular part of internal capsule mean diusivities | 6.06E-05 | 17706 | C | T |
| doi.org/10.11 01/288660 | 2019 | Neurological | Superior corona radiata radial diusivities | 6.76E-05 | 17706 | C | T |
| 31676860 | 2019 | Neurological | White matter | 7.26E-05 | 19629 | C | T |
| 31427789 | 2019 | Activities | Medication for cholesterol, blood pressure, diabetes, or take exogenous hormones: Cholesterol lowering medication | 7.47E-05 | 207533 | C | T |

**Chr19 Significant P-value 5.1E-04 rs612285**

| PMID | Year | Domain | Trait | P-value | N | EA | NEA |
|---|---|---|---|---|---|---|---|
| 30804566 | 2019 | Neurological | Frequent insomnia symptoms | 1.40E-04 | 237627 | A | G |
| 31427789 | 2019 | Reproduction | Age at menopause (last menstrual period) (female) | 2.89E-04 | 119160 | A | G |
| 31427789 | 2019 | Reproduction | Had menopause (female) | 3.35E-04 | 175519 | G | A |

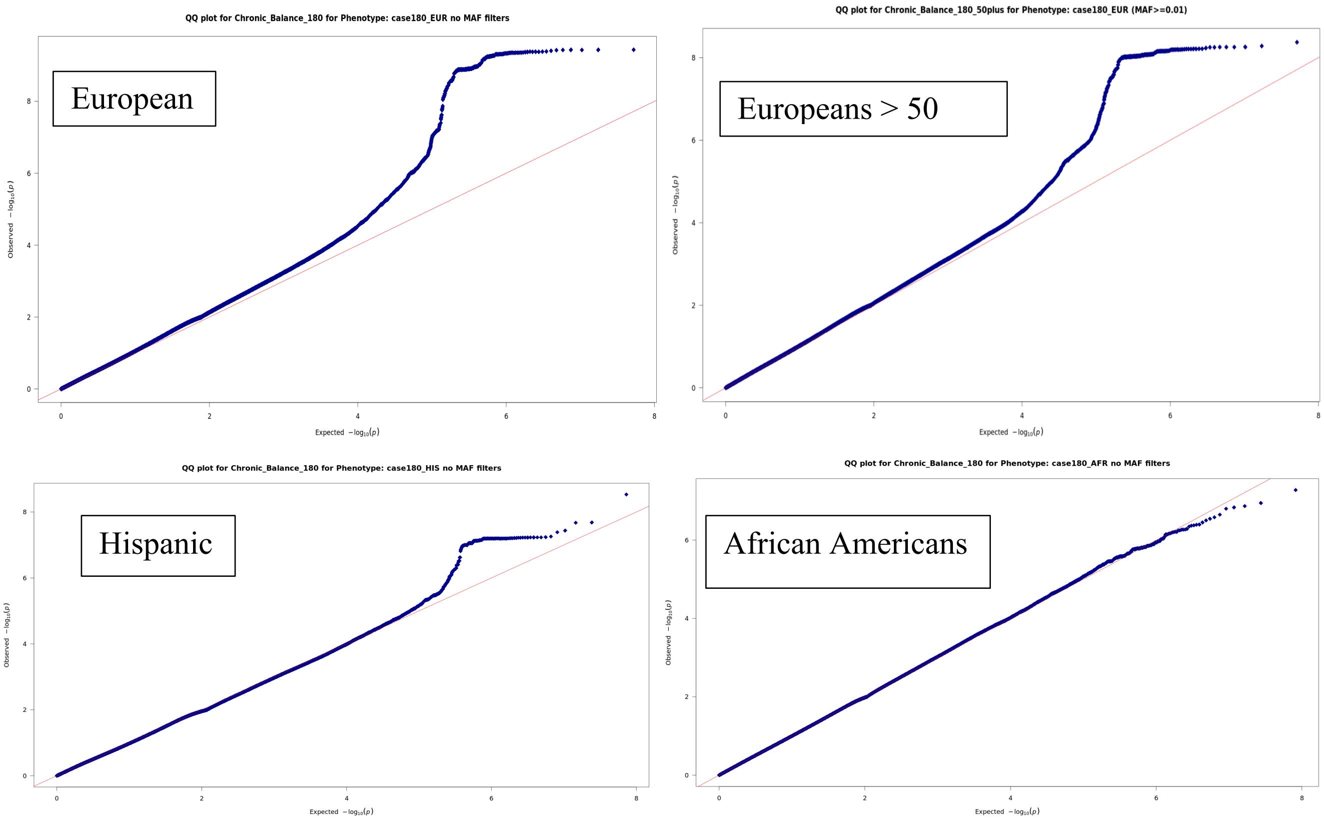


**Figure e1: Quantile-quantile plot of expected versus observed -log10 *P*-values for a genome-wide association study (GWAS) of balance.**

**A.** QQ-Plot European. **B.** QQ-Plot Europeans > 50 years of age**.** **C.** QQ-Plot Hispanic Ancestry. **D.** QQ-Plot African American Ancestry. Quantile-quantile plots show the distribution of expected versus observed *P* Values for the GWAS in the Million Veteran Program cohort consisting of 50,339 cases of chronic imbalance and 366,900 controls. Filtering was performed to include only SNPs with P-value < 1 x $10^{-05}$; For the European analysis, GC ƛ = 1.127; LDSC intercept ($h_{2\ int}$ = 1.027, SE = 0.0079). Polygenic effects account for 94.8% of the observed inflation.

**Figures e2 through e9. Chromosome base positions are aligned on each figure.**

**A.** Chromosomal position is indicated on the x-axis. -log10 *P*-values for each single nucleotide polymorphism (SNP) (filled circles) are indicated on the y-axis, with the lead SNP shown in purple. Annotated genes in the region are drawn in the lower panel. Recombination rate is indicated by a blue line. Additional SNPs in the locus are colored according to linkage disequilibrium ($r^2$) with the lead SNP. Recombination rates estimated from the CEU HapMap population represented by the vertical blue line.

**B.** Combined Annotated Dependent Depletion (CADD) scores for top, lead, and independent significant SNPs. CADD is an *in silico* prediction of variant pathogenicity. Scaled CADD score of 10 indicates that the variant is among the top 10% of deleterious variants.

**C.** RegulomeDb is a database with annotations of regulatory elements, i.e., transcription factor binding, transcription factor motifs, Dnase peaks and eQTLs. Lower numbers (1a-f) indicate a higher chance of a regulatory b binding site SNP. 7 indicates no binding site noted.

**D**. Chromatin state in the region of significant hits.

**E.** Where noted, eQTLs for genes within the locus, different colors represent different tissues that are indicated on the right side of the diagram. Significance is at FDR < .05.

**F.** Where noted, Hi-C results indicate intrachromatin capture (eFigure 9 only).

**Figure e2. Regional association plot (Locus zoom) top hit rs12779865 in trans-ancestry analysis.**

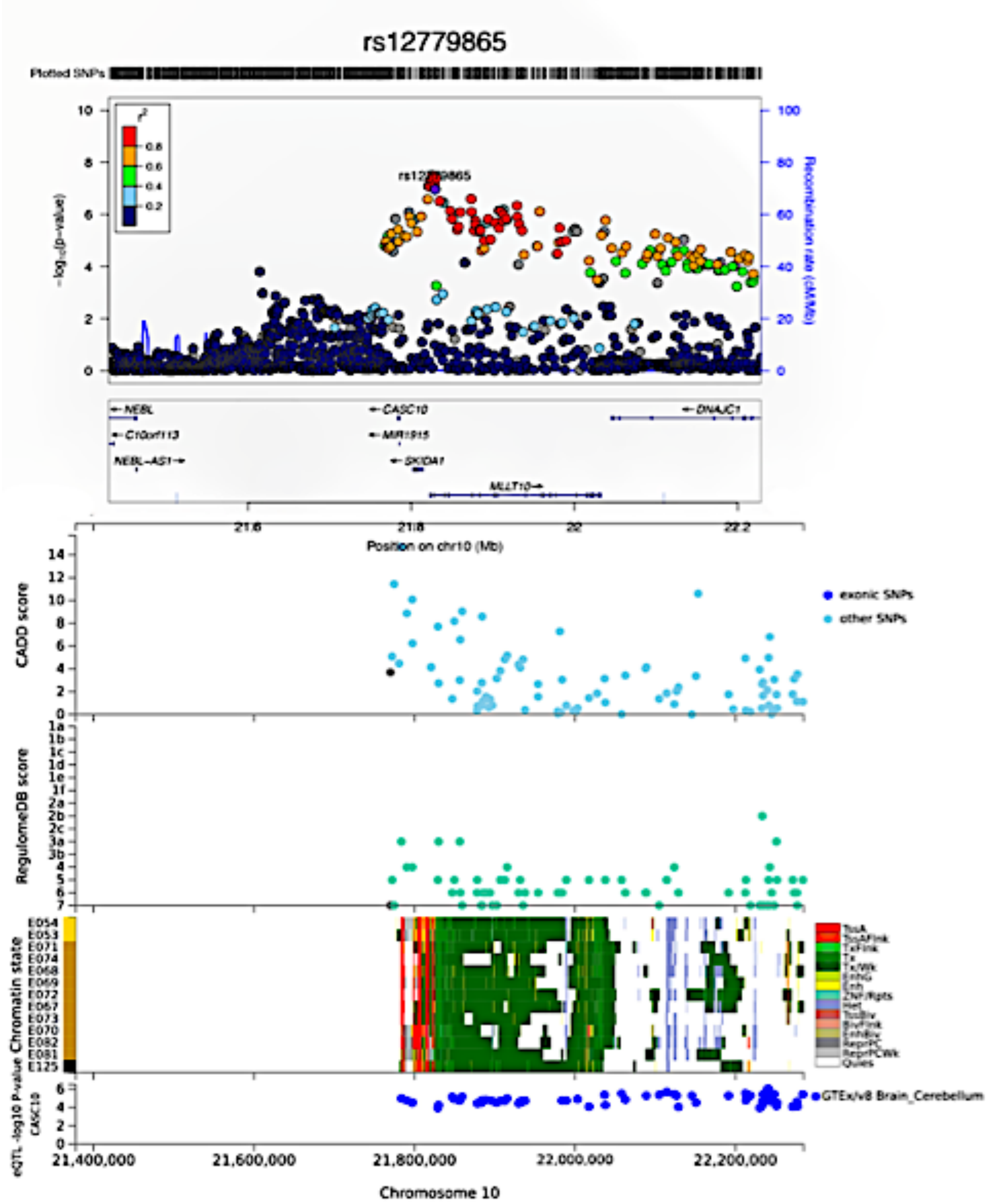

**Figure e3. Regional association plot (Locus zoom) top hit rs72631329 in trans-ancestry analysis.**

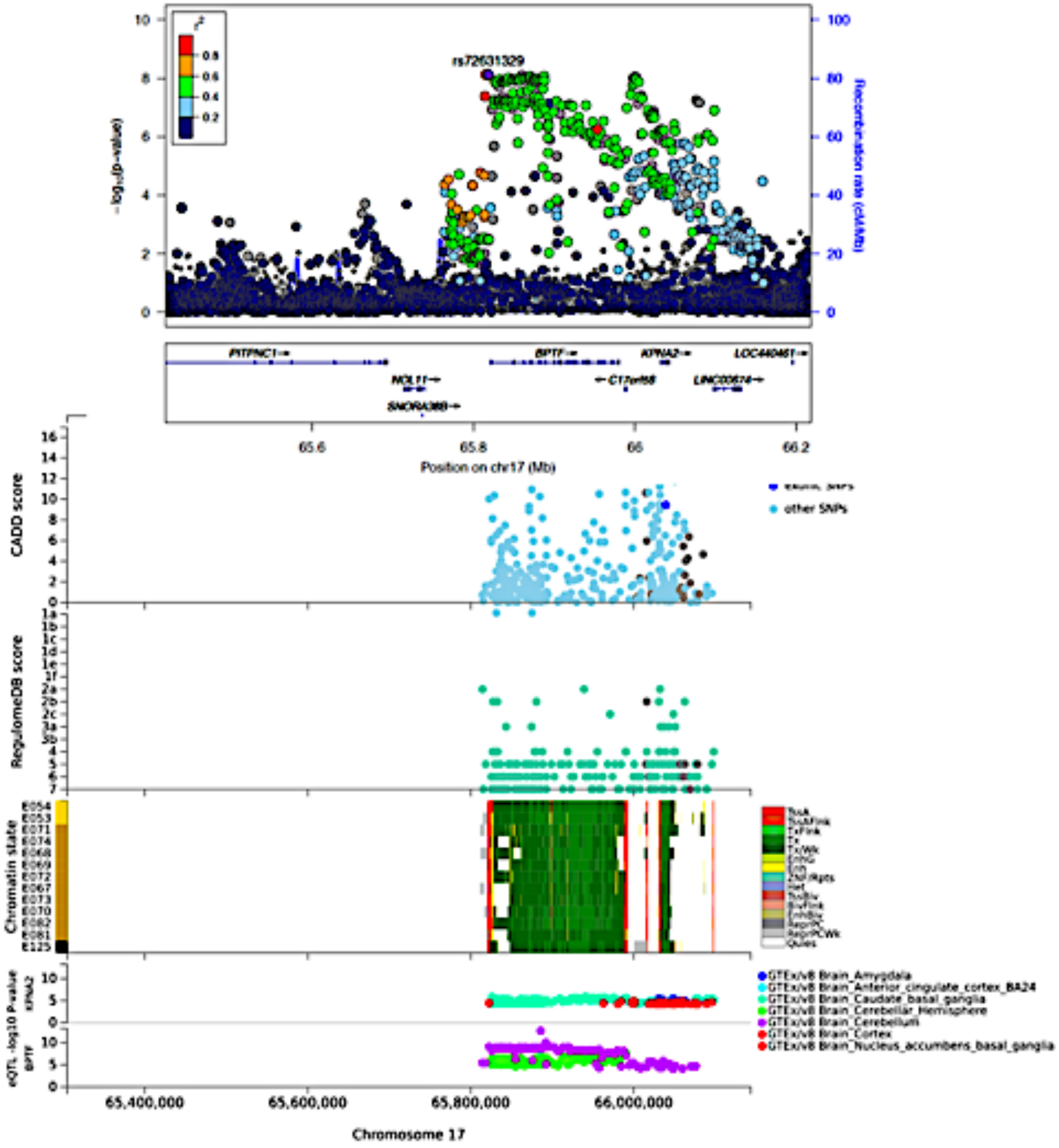

**Figure e4. Regional association plots (Locus zoom) top hit rs10713223 in trans-ancestry analysis.**

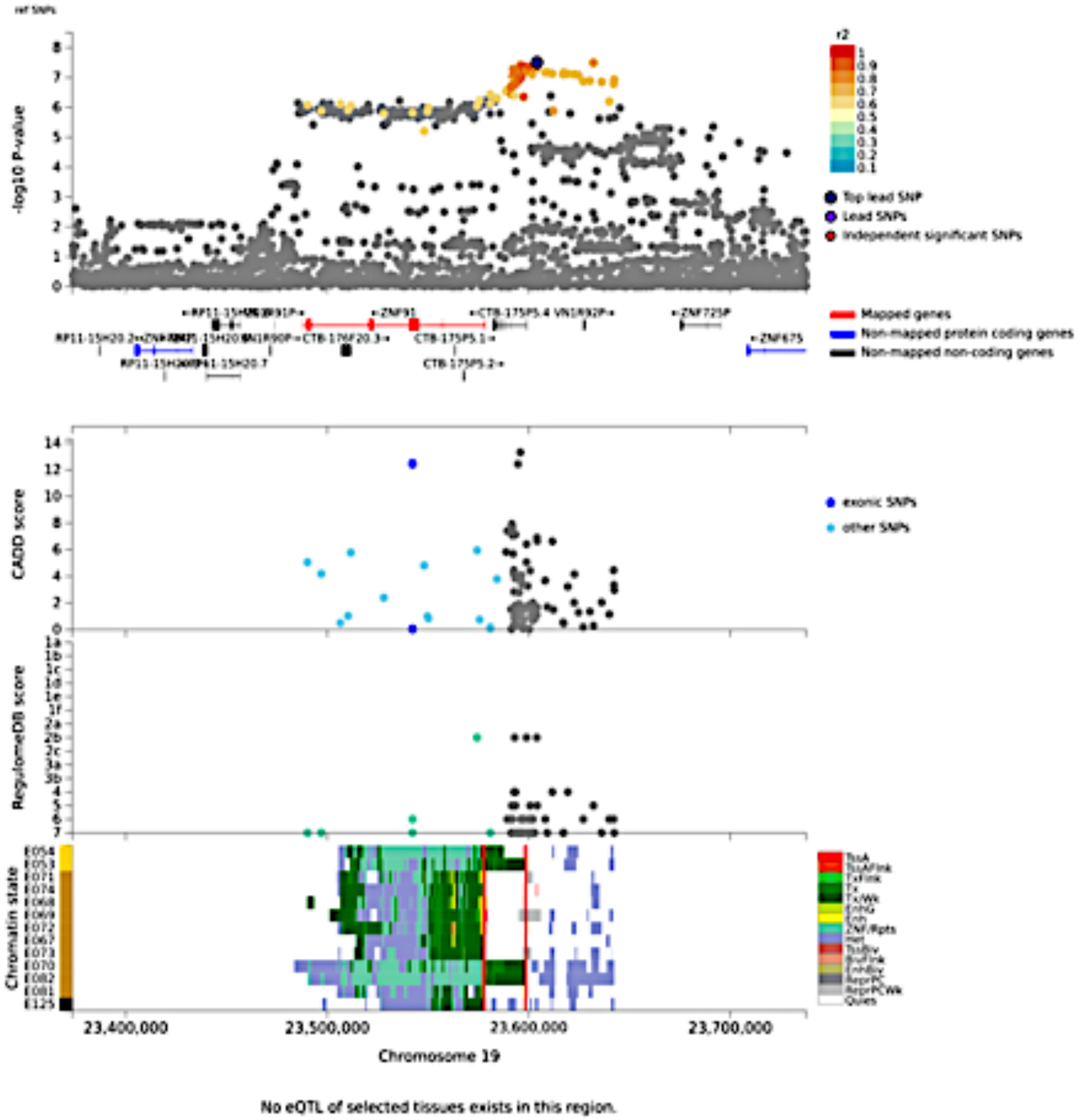

**Figure e5. Regional association plots (Locus zoom) of top hit rs200689072 in transancestry GWAS.**

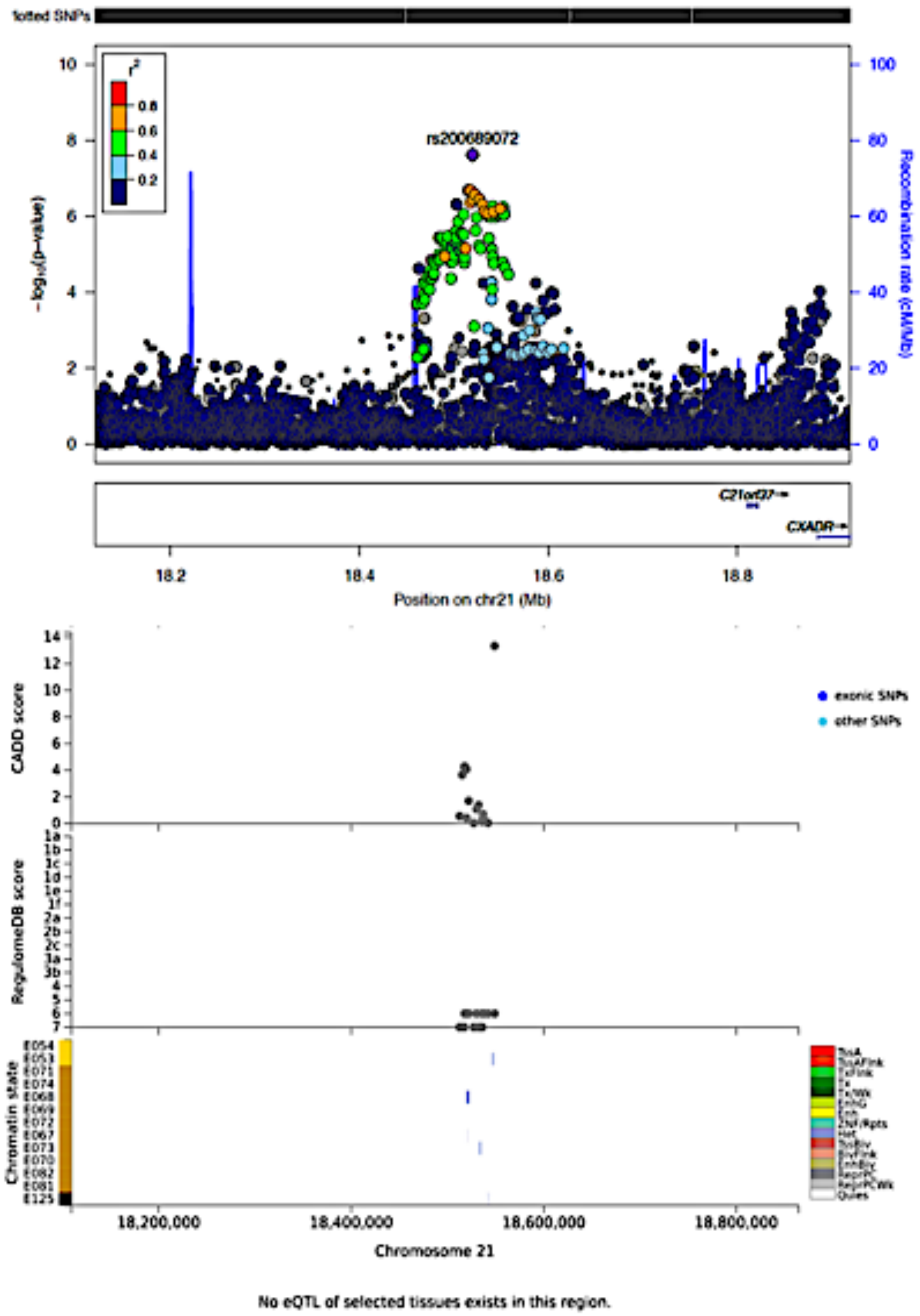

**eFigure 6. Regional association plots (Locus zoom) of top hit rs10828248 in European analysis.**

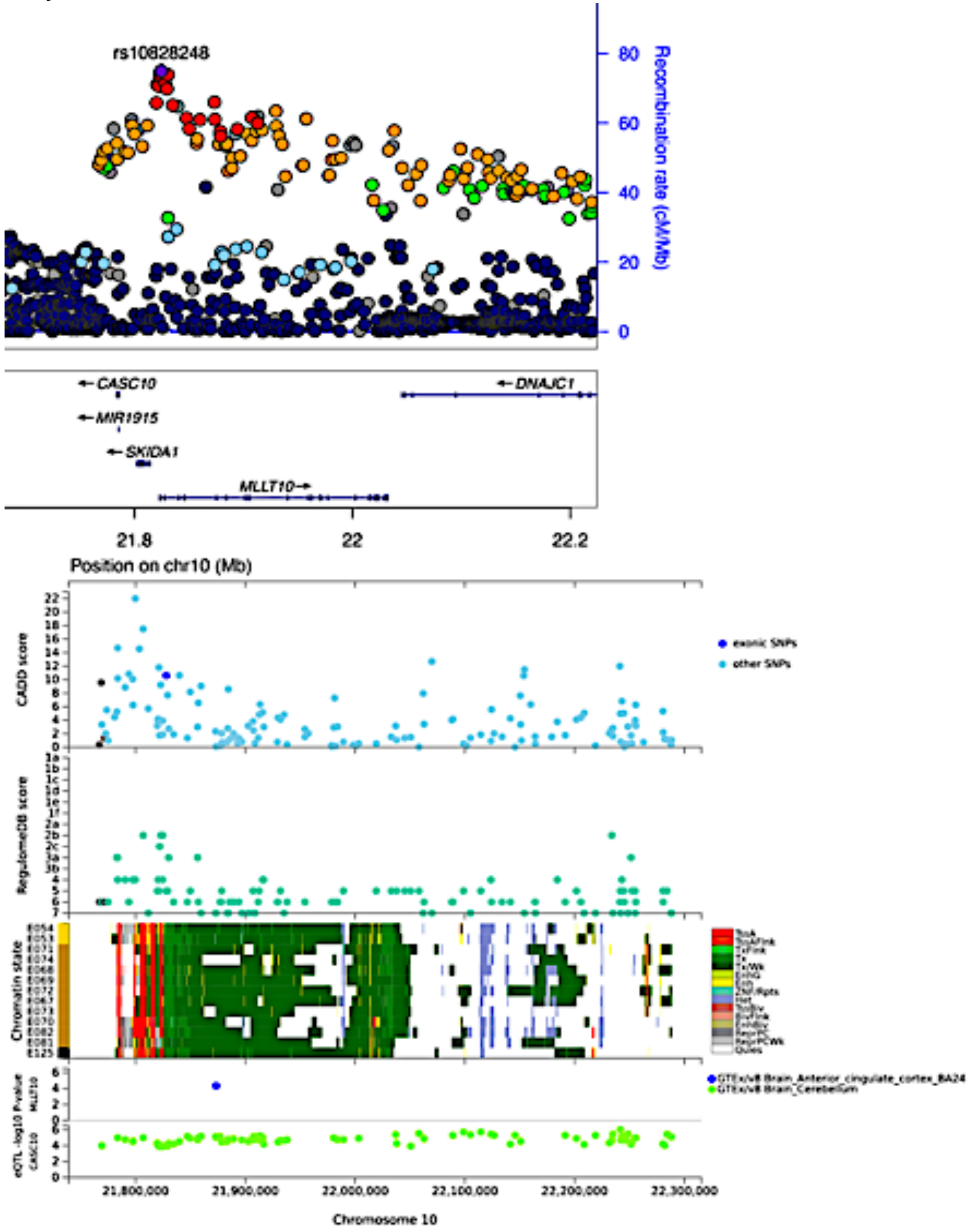

**eFigure 7. Regional association plots (Locus zoom) of top hit rs7221651 in European analysis.**

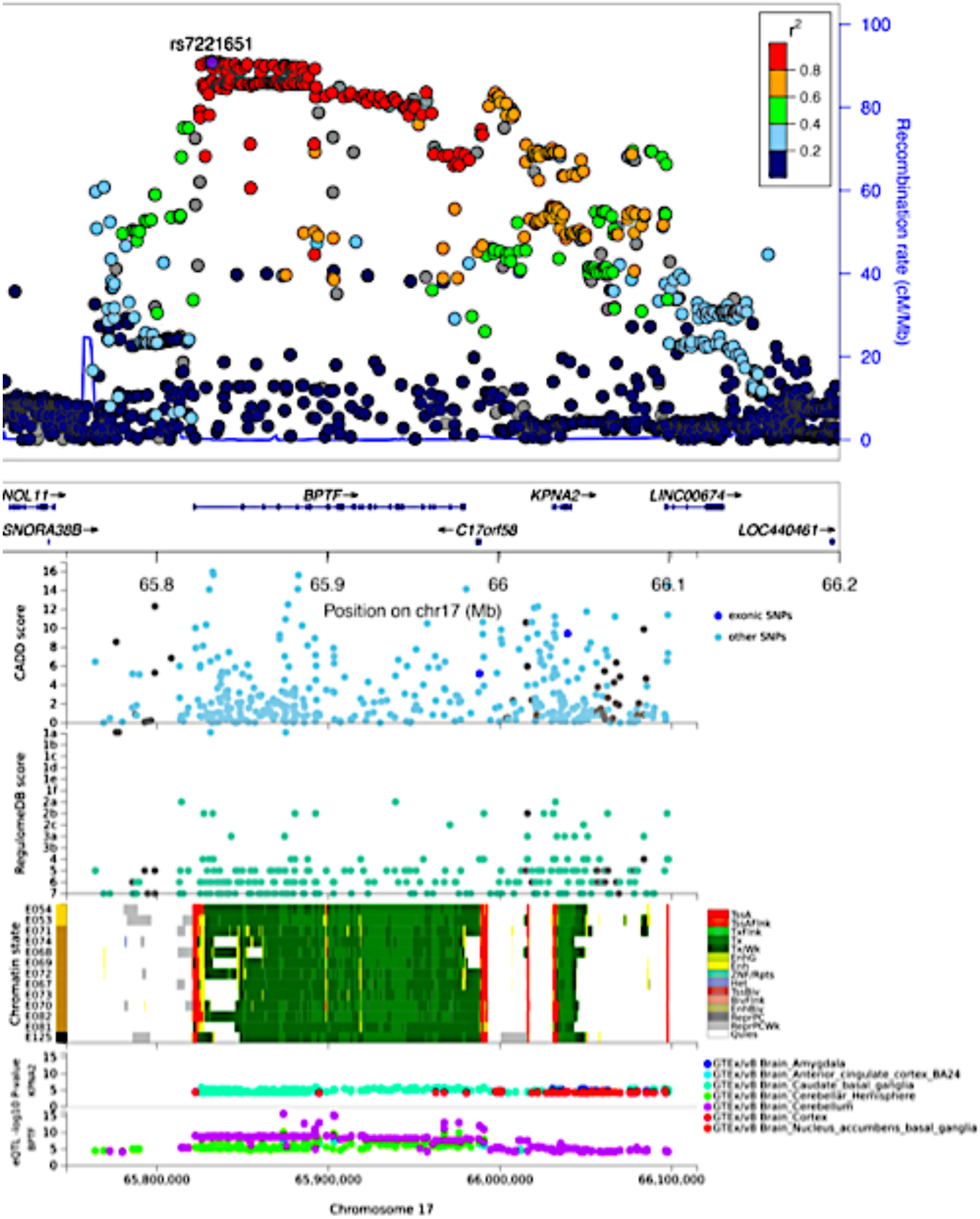

**eFigure 8. Regional association plots (Locus zoom) of top hit rs4791046 in Europeans over the age of 50.**

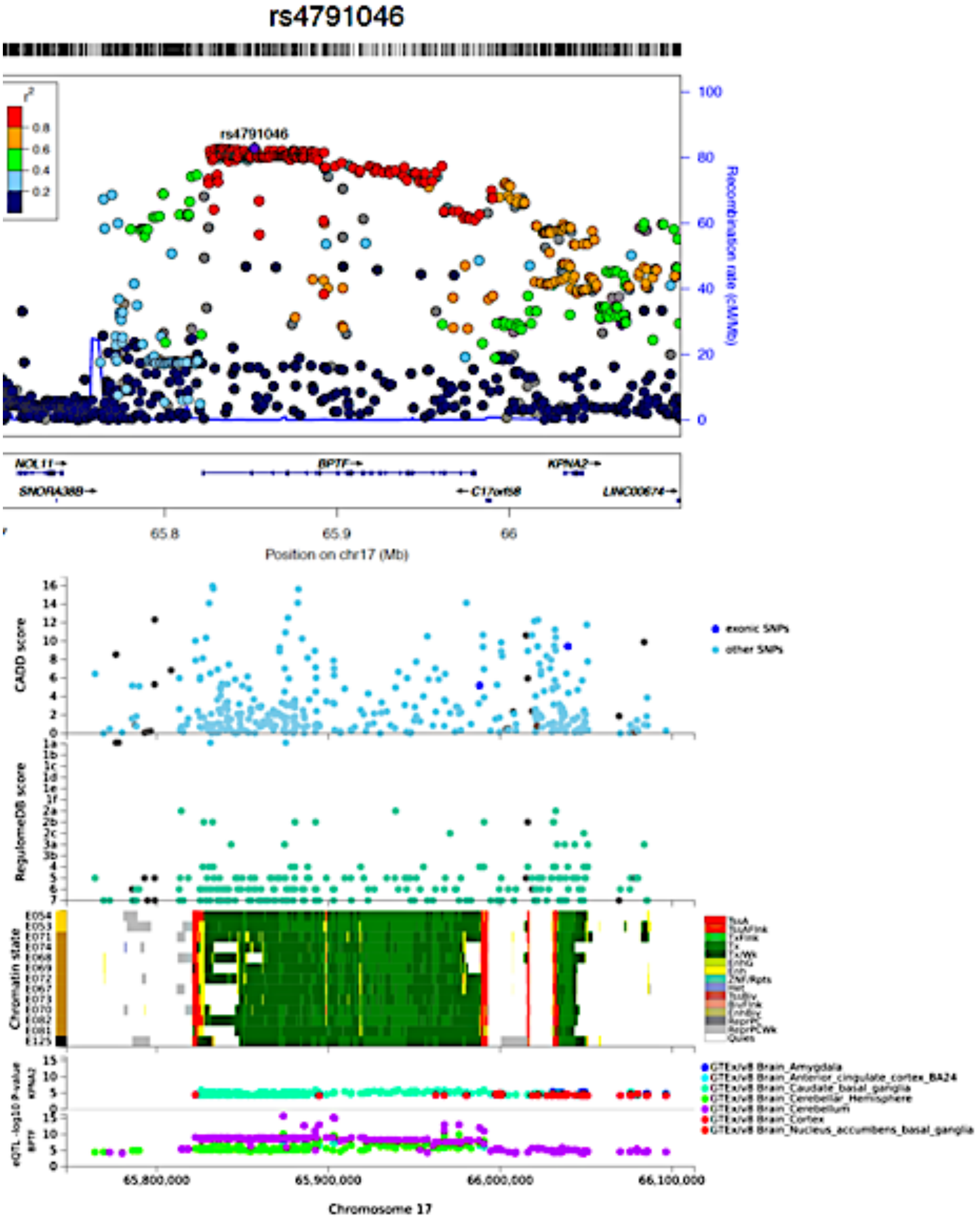

**eFigure 9. Regional plot of top hit rs71717606 in Hispanic ancestry GWAS.**

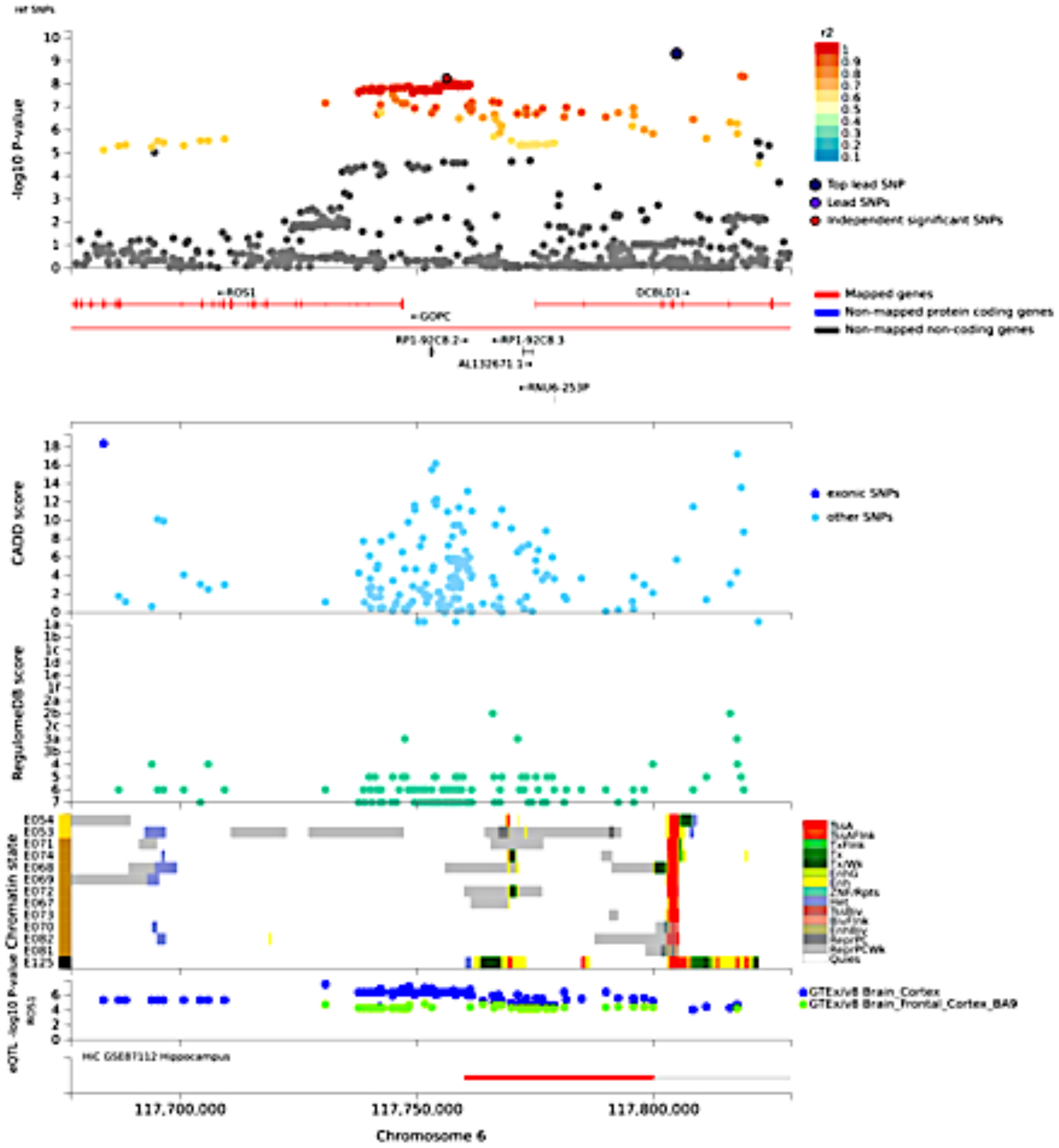

#### eFigure 10. Fine Mapping Plots

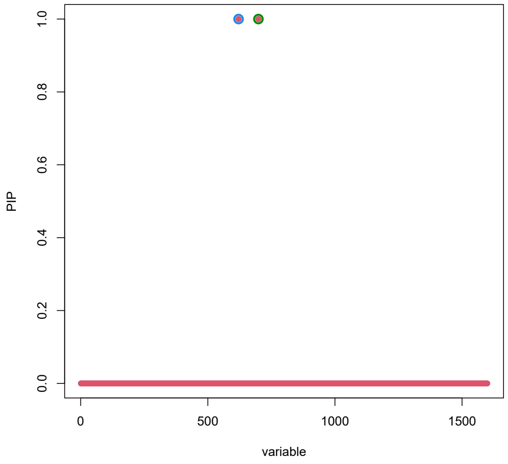


**a. Chromosome 6 – MXL ancestry**

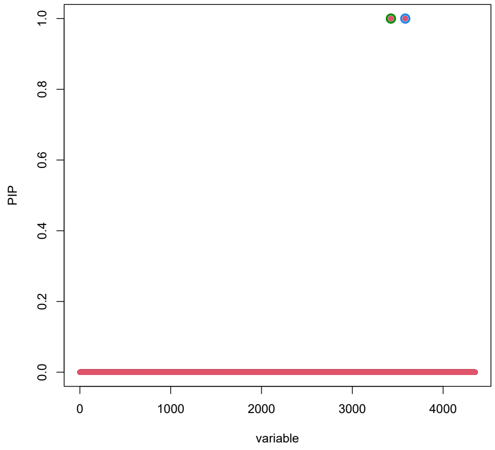


**b. Chromosome 10 – CEU ancestry**

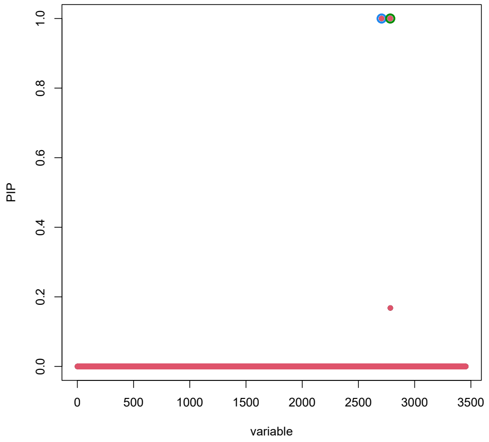


**c. Chromosome 17 – CEU ancestry**

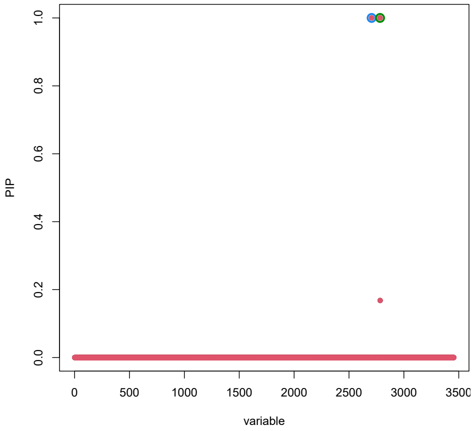


**d. Chromosome 19 – CEU, MXL, ASW**

**Notes: the red dots indicate SNPs that are part of credible sets. PIP indicates posterior inclusion probability of potential causal variants.**

#### Figure e11. PheWAS on lead SNPs

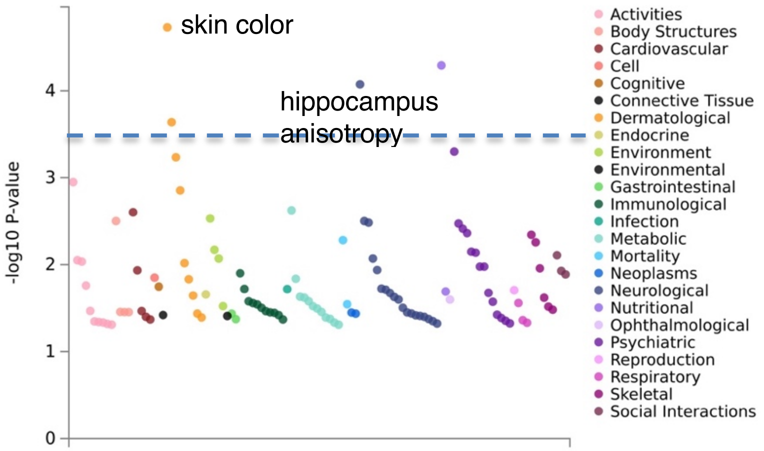


**Figure e11a. PheWAS chr6 - rs72965321 - on 3,300 traits. See Table e5 for numeric values. Dotted line indicates significant P-value < 2.94E-04.**

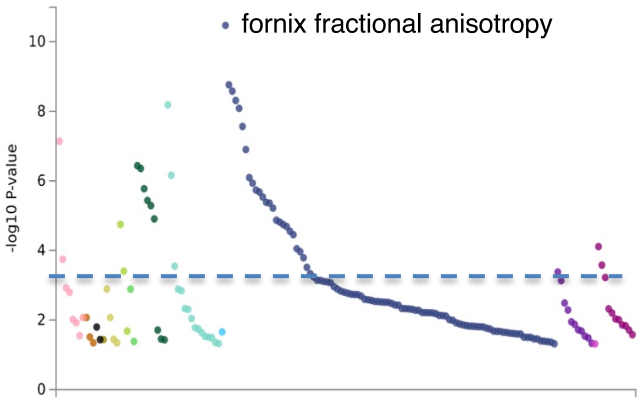


**Figure e11c. PheWAS chr10 - rs7894565 - on 3,300 traits. See Table e5 for numeric values. Dotted line indicates significant P-value < 4.31E-04.**

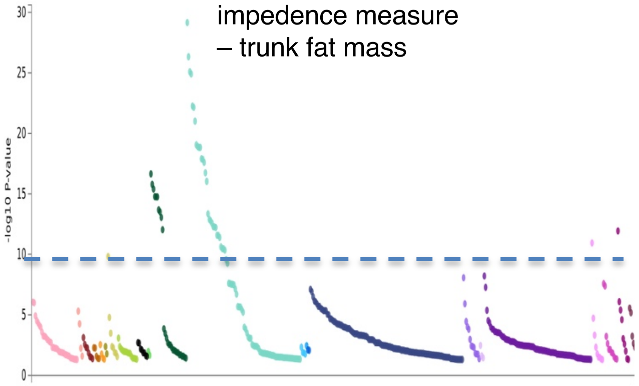


**Figure e11c. PheWAS chr17 - rs7221651 - on 3,300 traits. See Table e5 for numeric values. Dotted line indicates significant P-value < 8.98E-05.**

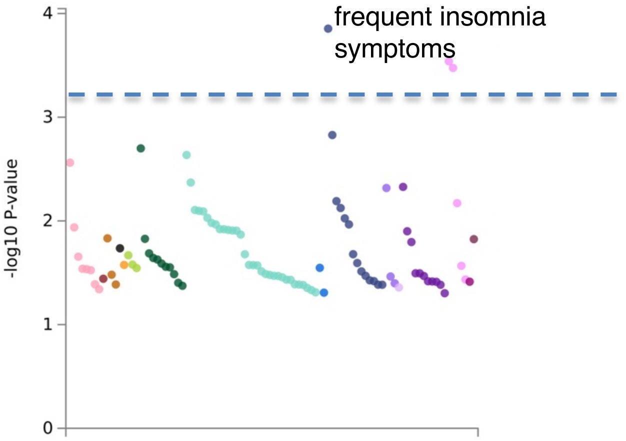


**Figure e11d. PheWAS chr19 - rs612285 - on 3,300 traits. See Table e5 for numeric values. Dotted line indicates significant P-value < 5.10E-04.**
